# Genetically Predicted Blood Metabolites Mediate the Association Between Gut Microbiota and Childhood with obesity: A Mendelian Randomization Study

**DOI:** 10.1101/2024.10.22.24315950

**Authors:** Min Zhang, Wenjuan Yan

## Abstract

**Background:** Childhood with obesity is characterized by metabolic dysregulation and unique gut microbiota profiles. Nevertheless, the comprehensive understanding of gut microbiota and metabolic dysregulation of Childhood with obesity remains unclear.

**Objectives:** This study aimed to investigate the causal relationship of gut microbiota and Childhood with obesity and identify the blood metabolites as potential mediators.

**Methods:** The exposure genome-wide association studies (GWAS) data were sourced from the GWAS Catalog, while the outcome GWAS data were obtained from the Early Growth Genetics (EGG) Consortium. The study used 473 types of gut microbiota, 233 types of blood metabolites, and Childhood with obesity from GWAS. We then performed two-sample Mendelian randomization (TSMR) and bidirectional Mendelian randomization (BDMR) analyses to explore the causal relationships between gut microbiota, blood metabolites, and Childhood with obesity. Additionally, we conducted multivariable Mendelian randomization (MVMR) and two-step Mendelian randomization (2SMR) to identify potential mediating blood metabolites in this process.

**Results:** MR analysis identified 13 types of gut microbiota and 12 types of blood metabolites that were causally associated with Childhood with obesity. Furthermore, there was no strong evidence that genetically predicted Childhood with obesity had an effect on these gut microbiota and blood metabolites. Further, 2SMR analysis revealed that the association between K10 sp001941205 and Childhood with obesity was mediated by the Total cholesterol to total lipids ratio in medium VLDL, accounting for 2.53% (95%CI; 2.14%-2.92%) of the association. Similarly, the relationship between SM23-33 and Childhood with obesity was mediated by the Ratio of 22:6 docosahexaenoic acid to total fatty acids, which accounted for 4.07% (95%CI; 2.70%-5.44%) of the association.

**Conclusions:** The present study is the first to investigate the causal relationships among 473 gut microbiota phenotypes, 233 blood metabolites, and Childhood with obesity through Mendelian randomization analysis, identifying 13 gut microbiota types with potential causal links to Childhood with obesity and suggesting that 2 blood metabolites may mediate these associations, thereby providing valuable insights for future intervention strategies aimed at addressing Childhood with obesity.

## 1 Introduction

Obesity is a long-term and complex condition where having too much body fat can seriously harm children’s health^1-2^. It can lead to a higher risk of type 2 diabetes^3^, and heart disease^4^, impact bone health,^5^ and reproductive functions,^6^ and even increase the likelihood of certain cancers^7^. Additionally, obesity can affect everyday activities like sleeping well and moving around comfortably.^8^ According to the latest figures from the World Health Organization, in 2022, approximately one in eight people worldwide was living with obesity. Since 1990, adult obesity rates have more than doubled, and the rates among teenagers have quadrupled. By 2022, there were over 390 million children and adolescents aged 5 to 19 years who were overweight, including 160 million classified as obese. Reducing the projected prevalence of overweight and obesity by 5% annually from current trends or maintaining it at 2019 levels would result in global economic savings of approximately US429 billion or US2201 billion per year, respectively, between 2020 and 2060.^9^ These statistics highlight the severity of the obesity epidemic and underscore the urgent need for effective strategies to combat this global health crisis.

The gut microbiota is crucial for digestion,^10^ immune regulation,^11^ and metabolism.^12^ A healthy gut microbiota is characterized by high diversity and stability;^13^ however, in obese children, microbial diversity is often lower.^14^ Dysbiosis of the gut microbiota may also trigger low-grade chronic inflammation, which is closely linked to insulin resistance and metabolic syndrome. These metabolic disorders further exacerbate the problem of obesity. Certain gut bacteria can enhance the host’s energy absorption. For example, the relative abundance of Firmicutes and Bacteroidetes is often more prevalent in obese children, suggesting that these microbial groups may promote fat accumulation by enhancing energy absorption efficiency in the host. ^15^ The gut microbiota produces metabolites through various metabolic pathways, and these metabolites can enter the circulatory system and have widespread effects on the host’s metabolism.Short-chain fatty acids (SCFAs), such as acetate, propionate, and butyrate, produced by the fermentation of dietary fiber by gut microbiota, can regulate energy metabolism, lipid metabolism, and glucose metabolism. SCFAs affect the host’s metabolic health through binding to G protein-coupled receptors (such as GPR41 and GPR43).^16^ Metabolites of the gut microbiota, such as lipopolysaccharides (LPS), are strong inflammatory mediators. LPS can enter the circulatory system by disrupting the intestinal barrier, triggering systemic inflammation, and thus promoting the onset of obesity-related metabolic diseases.^17^

Studying the relationship between Childhood with obesity, gut microbiota, and circulating metabolites is crucial for gaining a deeper understanding of the mechanisms underlying obesity, providing new perspectives for its prevention and treatment. Exploring the roles of gut microbiota and metabolites in obesity can offer a basis for developing new intervention strategies^18^. For example, regulating gut microbiota through probiotics and prebiotics may become an effective method for controlling Childhood with obesity.^19^ The involvement of gut microbiota and circulating metabolites in the onset and progression of Childhood with obesity is a complex and multifaceted process. An in-depth investigation of these mechanisms will not only help to understand the nature of obesity but also provide scientific evidence and new strategies for its prevention and treatment. While some progress has been made in research on the relationship between gut microbiota and Childhood with obesity, there are still significant gaps and pressing questions that need to be clarified. Many studies have found an association between gut microbiota and obesity,^20^ but in certain cases, the causal relationship remains unclear, particularly regarding whether changes in gut microbiota are a cause or a consequence of obesity in children.^21^ The composition of gut microbiota is highly complex, and some important but less abundant strains may be overlooked, necessitating more precise techniques to fully understand these symbiotic microorganisms. Additionally, gut microbiota exhibit significant individual variability, leading to substantial differences in microbial function and metabolic products among different individuals, which may limit the applicability of research findings.^22^

Mendelian randomization (MR) analysis is a robust epidemiological research strategy based on Mendelian inheritance. This technique allows for estimating causal relationships using genetic variants as instrumental variables, thereby addressing confounding variables, measurement errors, and reverse causation.^23^ We applied three large-scale genome-wide association studies (GWAS) to study the relationship between 473 gut microbiota and 233 blood metabolites and Childhood with obesity using MR. The aim is to elucidate the causal relationship between gut microbiota, blood metabolites, and Childhood with obesity, and to gain a clearer understanding of how gut microbiota affects Childhood with obesity through its effects on blood metabolites.

## 2 Materials and Methods

### 2.1 Study design

In this study, we employed a range of Mendelian Randomization (MR) methodologies, including two-sample MR (TSMR), bidirectional MR (BDMR), multivariable MR (MVMR), and two-step MR (2SMR), to investigate the causal relationships between gut microbiota, blood metabolites, and Childhood with obesity. By employing BDMR, we avoid the potential for reverse causation, ensuring that our findings accurately reflect the directionality of the relationships. We utilized MVMR to minimize bias and obtain reliable estimates of modifiable exposures and their relationships with targeted outcomes.

Our study design is based on three key assumptions essential for the causal interpretation of MR estimates. ^24^ We ensured the reliability of our findings by using genetic variations or single nucleotide polymorphisms (SNPs) as instrumental variables (IVs) that meet three crucial criteria: (i) the genetic IVs are strongly associated with exposure, (ii) the genetic IVs are not associated with confounders linked to the selected exposure and outcome, and (iii) the genetic IVs influence the outcome only through the exposure.25 Additionally, we completed the STROBE-MR checklist to ensure the integrity of this observational MR study;26 relevant details are provided in Supplementary Table S1, and a clear overview of our study design is presented in Figure 1.

**Figure 1.**
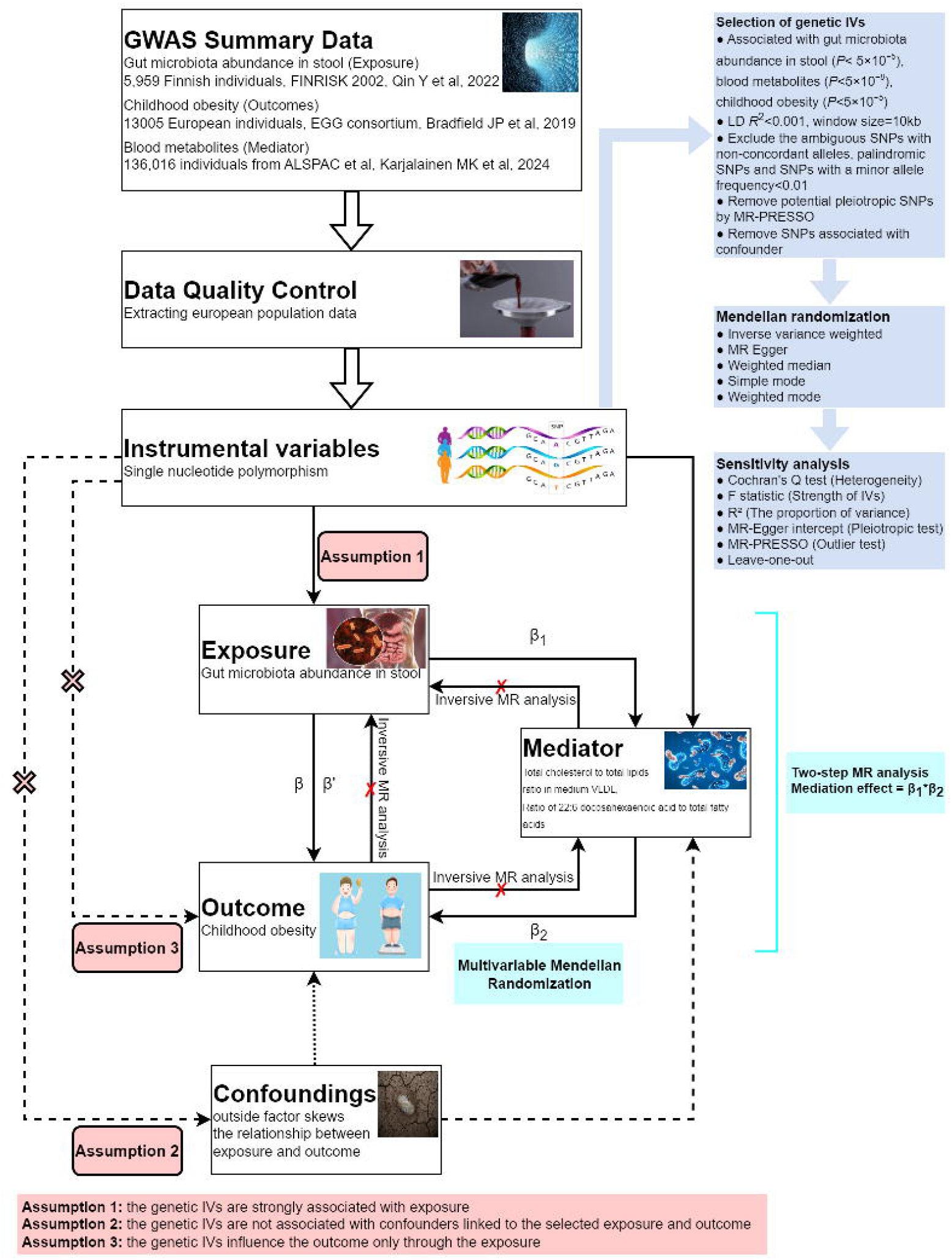
Overall study design plan. GWAS. genome-wide association studies: SNPs: single-nucleotide polymorphisms: β: the total causal effects of exposure on outcome was calculated using 2SMR method: β1 : the causal effects of exposure on mediators were calculated using 2SMR method. β2: the causal effects of mediators on outcome were calculated using MVMR method. β’: the direct causal effects of exposure on outcome were calculated using the formula β’=β-(β1×β2).

### 2.1 Data sources

Gut microbiota association studies (GWAS) data utilized in this research were obtained fr om a study of the genetic characteristics of gut microbiota. The original GWAS was cond ucted on 2,801 microbial taxa and 7,967,866 human genetic variants from 5,959 individu als enrolled in the FR02 cohort. GWAS summary data for Gut microbiota was downloade d from the GWAS Catalog (https://ftp.ebi.ac.uk/pub/databases/gwas/summary_statistics/), and the GWAS Catalog accession numbers range from GCST90032172 to GCST90032 644. More detailed information about the GWAS data can be obtained from their study.^27^

The GWAS data for blood metabolites were sourced from the GWAS Catalog, originally generated by Karjalainen MK et al.,^28^ with GWAS Catalog accession numbers ranging from GCST90301941 to GCST90302173. This comprehensive study identified genetic associations for 233 circulating metabolic traits across 33 cohorts, encompassing a total of 136,016 participants, predominantly of European ancestry, with a small proportion from Asia (11.60%,15775/136016).

Data on Childhood with obesity were contributed by the EGG consortium and downloaded from http://egg-consortium.org/childhood-obesity-2019.html. ^29^ To identify genetic variants associated with obesity in children, the researchers conducted a trans-ancestral meta-analysis of 30 studies, which included up to 13,005 cases and 15,599 controls from European, African, North American, South American, and East Asian ancestries. Following the screening, GWAS data specific to the European population were extracted for analysis.

### 2.3 Selection of IVs and data harmonization

To adhere to the stringent criteria based on the three principal MR assumptions and to mitigate horizontal pleiotropy, only independent genome-wide significant single nucleotide polymorphisms (SNPs) were employed as instrumental variables (IVs) for the exposure. The IVs must be closely related to the exposure (gut microbiota, blood metabolites, Childhood with obesity), and SNPs significantly associated with the occurrence were selected at the whole-genome level (*P*<5×10^−8^, *r*^2^<0.001, window size=10kb), If there are too few SNPs Included, the inclusion criteria can be changed to (*P*<5×10^−5^ or *P*<1×10^−5^, *r*^2^<0.001, window size=10kb). Additionally, we calculated the *F* statistics of the IVs to assess the extent of weak instrument bias. To reduce the bias caused by weak working variables, the working variables with *F* > 10 are retained. The formula for calculating the *F* value and *R*^2^ is as follows.

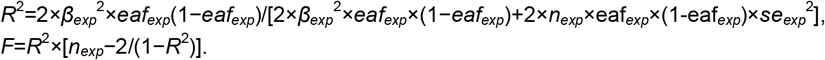

Finally, after data filtering, horizontal pleiotropy analysis was conducted. If there is horizontal pleiotropy, the MR-PRESSO test is performed. The outliers obtained from the test were removed before proceeding with further analysis.

### 2.4 Causal effects of gut microbiota on Childhood with obesity

We performed bidirectional MR analyses to investigate the causal relationship between gut microbiota and Childhood with obesity, estimating the total effect (β) of this relationship. We used the inverse variance weighted (IVW) method to estimate effects, reporting β±*SE* for continuous outcomes and *OR* (95%*CI*) for binary outcomes. In brief, the IVW method meta-analyzed SNP-specific Wald estimates by dividing the SNP-outcome association by the SNP-exposure association, using random effects to derive the final causal effect estimate. Additionally, we used MR-Egger and weighted median methods as complementary approaches to IVW, providing a more comprehensive understanding of the causal relationship.

### 2.5 Mediation analyses link “gut microbiota–blood metabolite–Childhood with obesity”

We further performed a mediation analysis using a two-step MR study to explore whether blood metabolite mediate the causal gut microbiota to Childhood with obesity.^30^ The specific approach is: (step 1) a two-sample MR (TSMR) model was carried out to estimate the effect of the exposure on the mediator, and (step 2) a second model estimating the effect of each mediator on the outcome was carried out using MVMR. Both the genetic variants for the mediator and the exposure were included in the first and second-stage regressions in MVMR. Using MVMR ensures that the mediator’s effect on the outcome is independent of the exposure. Additionally, this method provides an estimate of the direct effect of the exposure on the outcome. The two regression estimates from the second stage regression are multiplied together to estimate the indirect effect**^Error! Bookmark not defined^**..

With respect to our research, the overall effect can be decomposed into a direct effect (without mediators) and an indirect effect (through mediators). The total effect of gut microbiota on Childhood with obesity was decomposed: direct effects of gut microbiota on Childhood with obesity (overall effect) and indirect effects mediated by gut microbiota through the blood metabolite (mediation effect). The mediation effect was calculated through **β**_1_×**β**_2_ :(i) the causal effect of the mediator (blood metabolite) on the outcome (Childhood with obesity) adjusted for exposure-induced confounding (*β*_2_) and (ii) the causal effect of the exposure (gut microbiota) on the mediator (*β*_1_). We calculated the percentage mediated by the mediating effect by dividing the indirect effect by the total effect. Meanwhile, 95%CI was calculated using the delta or difference method.

### 2.6 Sensitivity analysis

The directional association between each identified SNP and both the exposure and outcome variables was assessed using MR Steiger filtering. This method measures the degree to which the variation in exposure and outcomes can be attributed to instrumental SNPs and determines if the variability in outcomes is less than that in exposure. Horizontal pleiotropy was further investigated via the MR-Egger approach, which utilizes weighted linear regression with an unconstrained intercept. This intercept acts as an indicator of the average pleiotropic effect across genetic variations, reflecting the typical direct influence of a variant on the outcome variable. If the intercept significantly deviated from zero (MR-Egger intercept *P*<0.05), it indicated the presence of horizontal pleiotropy. Moreover, Cochrane’s Q-test was used to assess heterogeneity, with lower p-values suggesting increased heterogeneity and a higher probability of directional pleiotropy. Leave-one-out analyses were also performed to identify potential SNP outliers.

### 2.7 Statistical analysis

We first used the results calculated by the IVW method as the final main result, and a significance threshold of *P*<0.05 was applied to the MR analysis, where P-values below this threshold were considered statistically significant. All statistical analyses and data visualizations were performed using R software (R Foundation, Vienna, Austria), with the TwoSampleMR (https://github.com/MRCIEU/TwoSampleMR) package for 2SMR and MVMR analysis and the PNG (Boutell, Netherlands) package for data visualization.

## 3 RESULTS

### 3.1 Selection of IVs

After screening, there were 13 different types of gut microbiota and 12 different types of blood metabolites with potential causal relationships with Childhood with obesity. The F-statistics for all IVs were above 10, indicating no evidence of weak instrument bias.^31^ After the Bonferroni adjustment, the p-values were all below the Bonferroni threshold.

### 3.2 Causal association between gut microbiota and Childhood with obesity

When evaluating the causal association of gut microbiota with Childhood with obesity, 13 types of gut microbiota show potential causal relationships. Relevant details can be found in Supplementary Table S2. Among them, the abundances in stool of SM23-33 (*OR=*0.755; 95%*CI=*0.599-0.951; *p=*0.017), TMED109 (*OR=*0.793; 95%*CI=*0.654-0.961; *p=*0.018), Eubacterium I ramulus A (*OR=*0.846; 95%*CI=*0.743-0.963; *p=*0.011), K10 sp001941205 (*OR=*0.895; 95%*CI=*0.805-0.997; *p=*0.043), Coprobacillus (*OR=*0.901; 95%*CI=*0.822-0.987; *p=*0.025), Ruminococcus E sp900314705 (*OR=*0.913; 95%*CI=*0.836-0.998; *p=*0.046), UBA1206 sp000433115 (*OR=*0.926; 95%*CI=*0.859-0.998; *p=*0.043), and Bacteroides A (*OR=*0.932; 95%*CI=*0.873-0.994; *p=*0.033) show a negative association with Childhood with obesity. This indicates that an increase in the abundance of these microbes leads to a decreased risk of Childhood with obesity.

Conversely, the following types of gut microbiota show a positive association with Childhood with obesity: Clostridium saudiense (*OR=*1.084; 95%*CI=*1.010-1.163; *p=*0.025), An181 (*OR=*1.151; 95%*CI=*1.013-1.307; *p=*0.030), Faecalicatena sp001517425 (*OR=*1.242; 95%*CI=*1.053-1.465; *p=*0.010), CAG-698 (*OR=*1.250; 95%*CI=*1.091-1.433; *p=*0.001), UBA8621 (*OR=*1.254; 95%*CI=*1.022-1.538; *p=*0.030), and Halomonadaceae (*OR=*1.499; 95%*CI=*1.005-2.236; *p=*0.047).

When evaluating the causal effects of Childhood with obesity on the gut microbiota, it was observed that all p-values were greater than 0.05, suggesting that Childhood with obesity does not have an effect on the gut microbiota under consideration. The relevant details are presented in Supplementary Table S3. The final analysis reveals potential causal relationships between 13 types of gut microbiota and Childhood with obesity, as illustrated in Figure 2.

**Figure 2.**
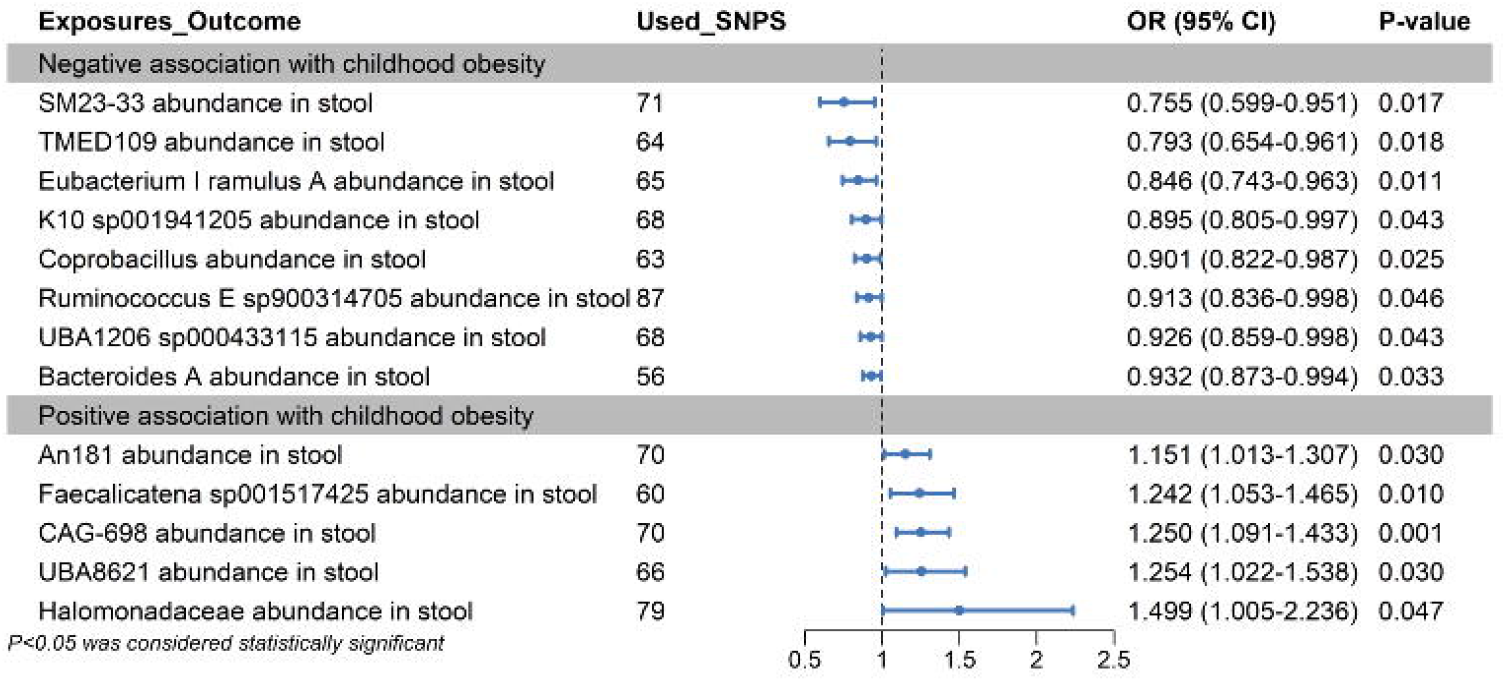
Mendelian randomization analyses show causal effects between gut microbiota and childhood with obesity. The forest plot shows significant causal associations with p<0.05 and the estimated odds ratio (OR) with 95% confidence intervals (CI).

### 3.3 Causal association between blood metabolite and Childhood with obesity

When evaluating the causal association of gut microbiota with Childhood with obesity, 12 types of blood metabolites show potential causal relationships. Relevant details are provided in Supplementary Table S4. Among them, the Triglycerides to total lipids ratio in medium VLDL shows a positive association with Childhood with obesity, indicating that an increase in Triglycerides to total lipids ratio in medium VLDL would lead to an increased risk of Childhood with obesity.

The other types of blood metabolites show a negative correlation. The IVW analysis results for these seven types of circulating immune cells are: Albumin levels(*OR=*0.717; 95%*CI=*0.524-0.981; *p=*0.037), Ratio of 22:6 docosahexaenoic acid to total fatty acids(*OR=*0.806; 95%*CI=*0.672-0.967; *p=*0.020), Total cholesterol levels in small HDL(*OR=*0.842; 95%*CI=*0.714-0.993; *p=*0.041), Cholesteryl esters to total lipids ratio in small HDL(*OR=*0.863; 95%*CI=*0.774-0.962; *p=*0.008), Total cholesterol to total lipids ratio in medium VLDL(*OR=*0.879; 95%*CI=*0.781-0.989; *p=*0.033), Total cholesterol to total lipids ratio in small HDL(*OR=*0.882; 95%*CI=*0.795-0.980; *p=*0.019), Omega-6 fatty acids levels(*OR=*0.883; 95%*CI=*0.790-0.986; *p=*0.028), Total lipids in IDL(*OR=*0.894; 95%*CI=*0.812-0.984; *p=*0.022), Total cholesterol to total lipids ratio in large LDL(*OR=*0.895; 95%*CI=*0.810-0.989; *p=*0.029), Phospholipids in very small VLDL(*OR=*0.900; 95%*CI=*0.824-0.984; *p=*0.021), Cholesteryl esters to total lipids ratio in small LDL(*OR=*0.902; 95%*CI=*0.817-0.996; *p=*0.042).

When evaluating the causal effects of Childhood with obesity on the blood metabolite, it was observed that all p-values were greater than 0.05, suggesting that Childhood with obesity does not have an effect on the gut microbiota under consideration. The relevant details are provided in Supplementary Table S5. The final analysis reveals potential causal relationships between 12 types of blood metabolites and Childhood with obesity, as illustrated in Figure 3.

**Figure 3.**
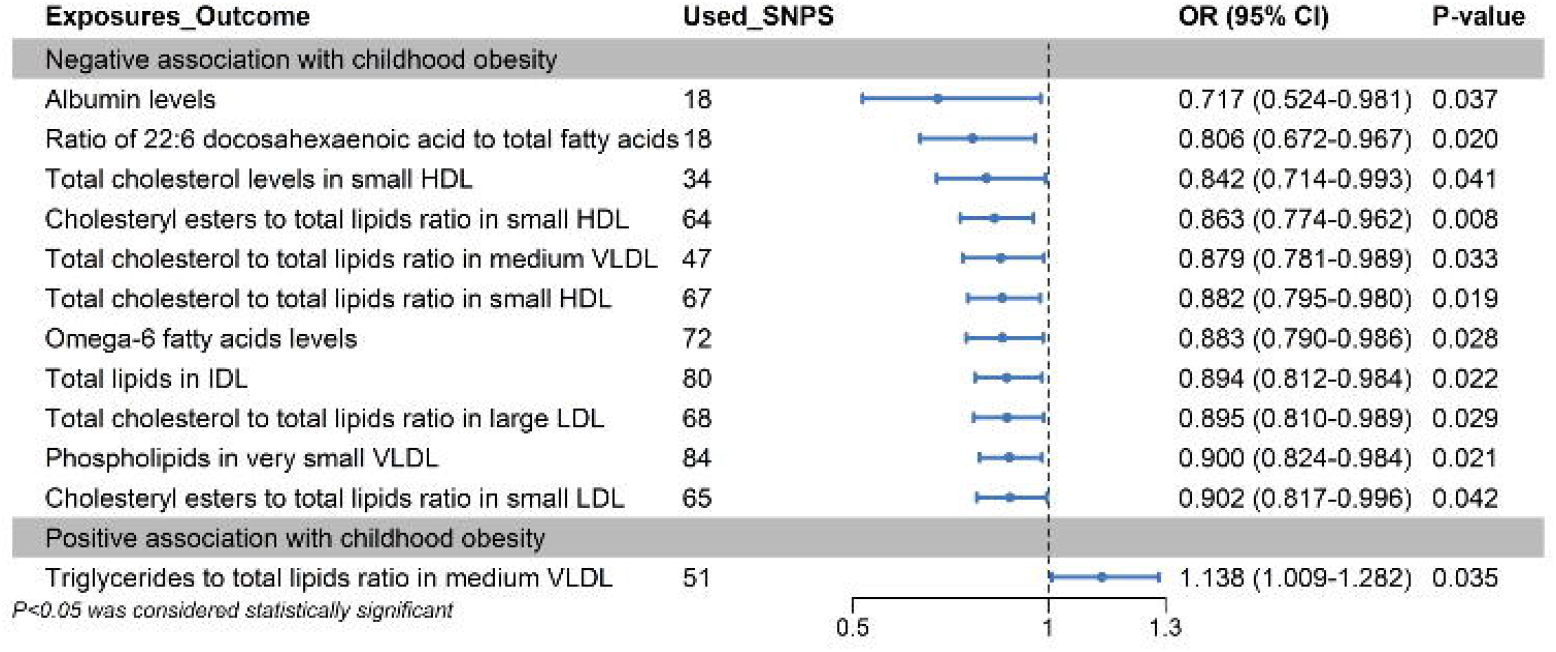
Mendelian randomization analyses show causal effects between blood metabolites and childhood with obesity. The forest plot shows significant causal associations with p<0.05 and the estimated odds ratio (OR) with 95% confidence intervals (CI).

### 3.4 Mediation MR analyses of blood metabolite

In the 2SMR analysis, 13 types of gut microbiota and 12 types of blood metabolites were found to be causally associated with Childhood with obesity, of which the abundance of K10 sp001941205 in stool was significantly associated with the Total cholesterol to total lipids ratio in medium VLDL. Similarly, the abundance of SM23-33 in stool was significantly associated with the ratio of 22:6 docosahexaenoic acid to total fatty acids. The results are presented in Table 1. According to all five methods, genetically predicted K10 sp001941205 abundance in stool was positively associated with the total cholesterol to total lipids ratio in medium VLDL (MR Egger; *OR=*1.010; 95%*CI=*0.966-1.057; *p=*0.654, Weighted median; *OR=*1.012; 95%*CI=*0.983-1.042; *p=*0.429, IVW; *OR=*1.024; 95%*CI=*1.003-1.045; *p=*0.024, Simple mode; *OR=*1.011; 95%*CI=*0.946-1.080; *p=*0.754, Weighted mode; *OR=*1.008; 95%*CI=*0.943-1.077; *p=*0.818). According to IVW methods, genetically predicted SM23-33 abundance in stool was positively associated with the ratio of 22:6 docosahexaenoic acid to total fatty acids (IVW; *OR=*1.059; 95%*CI=*1.013-1.108; *p=*0.012).

**Table 1.** Causal Effects Between Gut Microbiota, Blood Metabolites and Childhood with obesity.

| *method* | *nsnp* | *b* | *se* | *pval* | *lo_ci* | *up_ci* | *or* | *or_lci95* | *or_uci95* |
| --- | --- | --- | --- | --- | --- | --- | --- | --- | --- |
| K10 sp001941205 abundance in stool on Childhood with obesity |  |  |  |  |  |  |  |  |  |
| MR Egger | 68 | -0.110 | 0.129 | 0.400 | -0.363 | 0.144 | 0.896 | 0.695 | 1.155 |
| Weighted median | 68 | -0.087 | 0.074 | 0.239 | -0.232 | 0.058 | 0.916 | 0.793 | 1.060 |
| Inverse variance weighted | 68 | -0.110 | 0.055 | 0.043 | -0.217 | -0.003 | 0.895 | 0.805 | 0.997 |
| Simple mode | 68 | -0.041 | 0.175 | 0.816 | -0.383 | 0.302 | 0.960 | 0.682 | 1.352 |
| Weighted mode | 68 | -0.080 | 0.145 | 0.581 | -0.364 | 0.203 | 0.923 | 0.695 | 1.225 |
| Total cholesterol to total lipids ratio in medium VLDL on Childhood with obesity |  |  |  |  |  |  |  |  |  |
| MR Egger | 47 | -0.192 | 0.107 | 0.080 | -0.402 | 0.018 | 0.825 | 0.669 | 1.019 |
| Weighted median | 47 | -0.174 | 0.083 | 0.037 | -0.338 | -0.011 | 0.840 | 0.713 | 0.989 |
| Inverse variance weighted | 47 | -0.129 | 0.060 | 0.033 | -0.247 | -0.011 | 0.879 | 0.781 | 0.989 |
| Simple mode | 47 | -0.223 | 0.143 | 0.126 | -0.504 | 0.057 | 0.800 | 0.604 | 1.059 |
| Weighted mode | 47 | -0.214 | 0.085 | 0.015 | -0.380 | -0.047 | 0.807 | 0.684 | 0.954 |
| K10 sp001941205 abundance in stool on Total cholesterol to total lipids ratio in medium VLDL |  |  |  |  |  |  |  |  |  |
| MR Egger | 70 | 0.010 | 0.023 | 0.654 | -0.035 | 0.056 | 1.010 | 0.966 | 1.057 |
| Weighted median | 70 | 0.012 | 0.015 | 0.429 | -0.017 | 0.041 | 1.012 | 0.983 | 1.042 |
| Inverse variance weighted | 70 | 0.024 | 0.011 | 0.024 | 0.003 | 0.044 | 1.024 | 1.003 | 1.045 |
| Simple mode | 70 | 0.011 | 0.034 | 0.754 | -0.055 | 0.077 | 1.011 | 0.946 | 1.080 |
| Weighted mode | 70 | 0.008 | 0.034 | 0.818 | -0.058 | 0.074 | 1.008 | 0.943 | 1.077 |
| SM23-33 abundance in stool on Childhood with obesity |  |  |  |  |  |  |  |  |  |
| MR Egger | 71 | -0.261 | 0.304 | 0.393 | -0.857 | 0.335 | 0.770 | 0.425 | 1.397 |
| Weighted median | 71 | -0.142 | 0.169 | 0.400 | -0.473 | 0.189 | 0.868 | 0.623 | 1.208 |
| Inverse variance weighted | 71 | -0.282 | 0.118 | 0.017 | -0.513 | -0.050 | 0.755 | 0.599 | 0.951 |
| Simple mode | 71 | -0.080 | 0.387 | 0.837 | -0.839 | 0.679 | 0.923 | 0.432 | 1.972 |
| Weighted mode | 71 | -0.080 | 0.342 | 0.816 | -0.751 | 0.591 | 0.923 | 0.472 | 1.806 |
| Ratio of 22:6 docosahexaenoic acid to total fatty acids on Childhood with obesity |  |  |  |  |  |  |  |  |  |
| MR Egger | 18 | -0.206 | 0.138 | 0.155 | -0.476 | 0.065 | 0.814 | 0.621 | 1.067 |
| Weighted median | 18 | -0.185 | 0.088 | 0.035 | -0.357 | -0.013 | 0.831 | 0.700 | 0.987 |
| Inverse variance weighted | 18 | -0.215 | 0.093 | 0.020 | -0.397 | -0.033 | 0.806 | 0.672 | 0.967 |
| Simple mode | 18 | -0.073 | 0.221 | 0.744 | -0.506 | 0.359 | 0.929 | 0.603 | 1.432 |
| Weighted mode | 18 | -0.176 | 0.081 | 0.045 | -0.336 | -0.017 | 0.838 | 0.715 | 0.983 |
| SM23-33 abundance in stool on Ratio of 22:6 docosahexaenoic acid to total fatty acids |  |  |  |  |  |  |  |  |  |
| MR Egger | 71 | 0.110 | 0.053 | 0.044 | 0.005 | 0.215 | 1.116 | 1.005 | 1.239 |
| Weighted median | 71 | 0.055 | 0.033 | 0.095 | -0.010 | 0.120 | 1.057 | 0.990 | 1.128 |
| Inverse variance weighted | 71 | 0.057 | 0.023 | 0.012 | 0.012 | 0.102 | 1.059 | 1.013 | 1.108 |
| Simple mode | 71 | 0.039 | 0.080 | 0.627 | -0.118 | 0.197 | 1.040 | 0.888 | 1.218 |
| Weighted mode | 71 | 0.042 | 0.077 | 0.589 | -0.110 | 0.194 | 1.043 | 0.896 | 1.214 |

When evaluating the causal effects of blood metabolites on gut microbiota, it was observed that all p-values were greater than 0.05, suggesting that blood metabolites do not have an effect on the gut microbiota under consideration. The relevant details are provided in Supplementary Table S6.

We found that the abundance of K10 sp001941205 in stool was associated with an increased Total cholesterol to total lipids ratio in medium VLDL, and this increased ratio was associated with a decreased risk of Childhood with obesity. Our study showed that the Total cholesterol to total lipids ratio in medium VLDL accounted for 2.53% (95%CI; 2.14%-2.92%) of the decreased risk associated with K10 sp001941205 abundance in stool concerning Total cholesterol to total lipids ratio in medium VLDL and Childhood with obesity. These results preliminarily illustrate that K10 sp001941205 abundance in stool may reduce the risk of Childhood with obesity partially by enhancing the effects of the Total cholesterol to total lipids ratio in medium VLDL on Childhood with obesity.

In addition to these findings, we observed a consistent relationship among SM23-33 abundance in stool, the Ratio of 22:6 docosahexaenoic acid to total fatty acids, and Childhood with obesity. Unlike the previous association, the mediation effect of the Ratio of 22:6 docosahexaenoic acid to total fatty acids accounted for 4.07% (95%CI; 2.70%-5.44%). The results are presented in Figure 4. The details of the MVMR analysis are presented in Supplementary Table S7.

**Figure 4.**
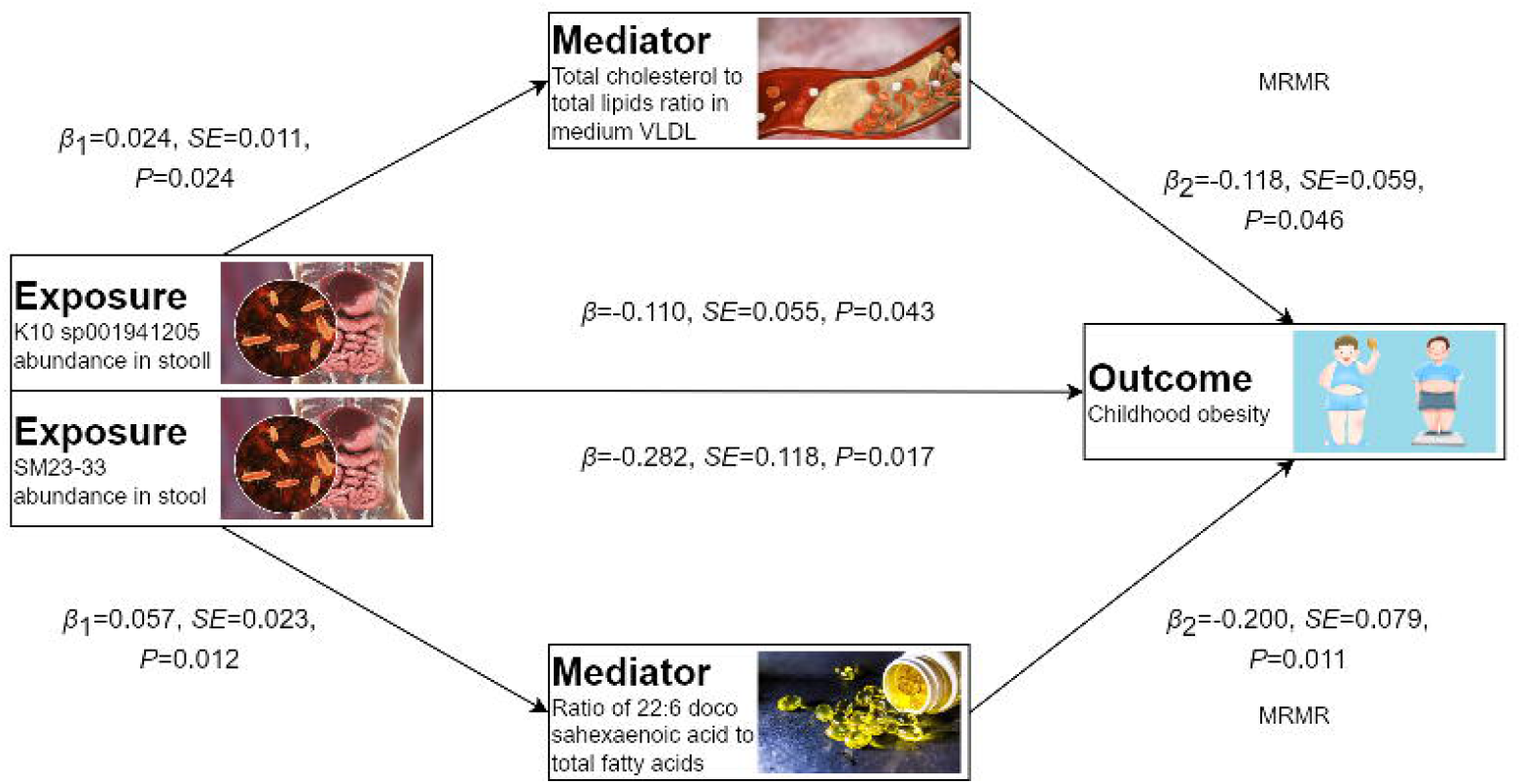
Schematic diagram of the mediating effect of blood metabolite between Gut microbiota and Childhood with obesity.

### 3.5 Sensitivity analysis

To address potential pleiotropy in our causal effect estimates, we performed multiple sensitivity analyses. Cochran’s Q-test and funnel plot analysis revealed no evidence of heterogeneity or asymmetry among the SNPs involved in the causal relationship, except for CAG-698 abundance in stool on Childhood with obesity (MR Egger *Q*=27.222; *df*=16; *p*=0.039, IVW *Q*=91.193; *df*=69; *p*=0.038) and Ratio of 22:6 docosahexaenoic acid to total fatty acids on Childhood with obesity (MR Egger *Q*=91.186; *df*=68; *p*=0.032). Furthermore, leave-one-out analysis validated the impact of each SNP on the overall causal estimates. Moreover, re-analysis of the MR study after excluding individual SNPs consistently yielded similar results, indicating that all SNPs contributed significantly to the establishment of the causal relationship, with relevant details provided in Supplementary Table S8.

## 4 Discussion

In the present study, we investigated the causal association among 473 gut microbiota phenotypes, 233 blood metabolites, and Childhood with obesity using large-scale genetic data and MR analysis. To our knowledge, this is the first MR analysis to explore the causal relationships among various gut microbiota phenotypes, blood metabolites, and Childhood with obesity. Through stringent inclusion criteria and sensitivity analyses, we have identified potential causal links between 13 distinct gut microbiota types and Childhood with obesity, with 2 specific blood metabolites potentially serving as mediators in this pathway.

Our study highlights significant associations between various types of gut microbiota and Childhood with obesity, contributing to the body of evidence regarding the role of the gut microbiome in Childhood with obesity. We identified 13 gut microbiota species with potential causal relationships to Childhood with obesity, which opens avenues for further exploration. Several taxa exhibited negative associations with Childhood with obesity. S Specifically, higher abundances in the stool of SM23-33, TMED109, Eubacterium I ramulus A, K10 sp001941205, Coprobacillus, Ruminococcus E sp900314705, UBA1206 sp000433115, and Bacteroides A were linked to a reduced risk of Childhood with obesity. This is consistent with findings from previous studies, ^32 - 33^ which emphasized the protective role of specific gut microbiota against metabolic diseases by promoting short-chain fatty acid production and enhancing gut barrier integrity.^34^ These microbes may play a protective role in Childhood with obesity, possibly through mechanisms such as improved metabolic regulation and enhanced satiety signaling. ^35^ Furthermore, a systematic review suggests that a diverse gut microbiota rich in beneficial species can offer resilience against obesity, further reinforcing our findings about the protective implications of specific bacterial populations^36^. Conversely, certain taxa demonstrated positive associations with Childhood with obesity, including Clostridium saudiense, An181, Faecalicatena sp001517425, CAG-698, UBA8621, and Halomonadaceae. These findings align with previous research, which identified that increased abundances of specific harmful bacteria could contribute to inflammation and insulin resistance, subsequently leading to obesity.^37^ The role of these microbes in promoting obesity raises important questions regarding their mechanisms, which may involve inflammatory pathways and metabolic disruption. ^38 - 39^ Our results emphasize the necessity of understanding the complex interplay between gut microbiota and host metabolism, encouraging future research to delve deeper into these relationships. Longitudinal studies that explore shifts in gut microbiota composition throughout childhood, as well as interventional studies targeting specific microbiota, will be crucial. These investigations could help identify effective strategies for obesity prevention and management through microbiota modulation. Recognizing these microbial profiles could inform future therapeutic strategies targeting gut health to combat the growing challenge of Childhood with obesity.

This study provides important insights into the causal relationships between gut microbiome-associated blood metabolites and Childhood with obesity. Using Mendelian randomization analysis, we identified 12 types of blood metabolites that show potential causal associations with the risk of Childhood with obesity. Notably, the Triglycerides to total lipids ratio in medium VLDL exhibited a positive causal relationship with Childhood with obesity. This indicates that an increase in this ratio would lead to an elevated risk of Childhood with obesity. This finding is consistent with previous evidence suggesting that higher triglyceride levels in VLDL are associated with an increased risk of obesity and metabolic disorders. In contrast, the other 11 blood metabolites showed inverse causal relationships with Childhood with obesity risk. These include Albumin levels, the Ratio of 22:6 docosahexaenoic acid (DHA) to total fatty acids, Total cholesterol levels in small HDL, Cholesteryl esters to total lipids ratio in small HDL, Total cholesterol to total lipids ratio in medium VLDL, Total cholesterol to total lipids ratio in small HDL, Omega-6 fatty acid levels, Total lipids in IDL, Total cholesterol to total lipids ratio in large LDL, Phospholipids in very small VLDL, and Cholesteryl esters to total lipids ratio in small LDL. These inverse causal associations suggest that higher levels or ratios of these metabolites may confer a protective effect against Childhood with obesity. For instance, the beneficial effects of DHA and omega-6 fatty acids on metabolic health and obesity risk have been well-documented.^40-41^ Additionally, the ratios of cholesterol to total lipids in various lipoprotein fractions may reflect favorable lipid profiles associated with a lower obesity risk.

This study has identified a significant relationship between the abundance of the gut microbiome species K10 sp001941205 and Total cholesterol to total lipids ratio in medium VLDL. Our findings indicate that this VLDL ratio can account for approximately 2.53% (95%CI; 2.14%-2.92%) of the decline in obesity risk associated with the abundance of K10 sp001941205. This suggests that K10 sp001941205 may partially reduce the risk of Childhood with obesity by enhancing the effects of the VLDL cholesterol to lipid ratio. Additionally, we observed a consistent association between the abundance of SM23-33, Ratio of 22:6 docosahexaenoic acid to total fatty acids, and Childhood with obesity. Unlike the prior relationship, the mediation effect of the DHA ratio accounted for 4.07% (95%CI; 2.70%-5.44%) of the decrease in Childhood with obesity risk. These findings enrich current literature by highlighting the role of gut microbiota in modulating lipid metabolism, which in turn influences obesity risk in children. Previous studies have suggested a negative relationship between triglycerides in VLDL and obesity risk.^42^ Conversely, our findings, suggest that an elevated VLDL cholesterol to lipid ratio may provide a protective effect, potentially due to the complex interrelations between these lipid components and obesity. Furthermore, increased DHA intake and its ratio to total fatty acids have been linked to improved metabolic health and a lower risk of obesity,^43-44^ aligning with our observations. These results indicate that gut microbiota may influence metabolic health and obesity outcomes by modulating specific blood metabolites. This microbiome-host interaction appears to be a complex dynamic process involving multiple mechanisms.^45^ Future research should delve deeper into how these key microbial species regulate lipid metabolism, potentially offering new targets for preventing and treating Childhood with obesity. Moreover, considering the high individual variability in metabolite and microbiome interactions, targeted interventions will be critical.^46-47^ In conclusion, this study highlights the complex association between specific gut bacteria, their metabolites, and the risk of Childhood with obesity. These insights provide a new perspective for understanding the mechanisms underlying metabolic diseases in children and lay a foundation for developing future prevention and treatment strategies. The identification of these causal relationships between gut microbiome-related blood metabolites and Childhood with obesity provides valuable insights into the underlying mechanisms. It highlights the complex interplay between the gut microbiome, lipid metabolism, and obesity development. These findings suggest that targeting specific microbial signatures and their associated metabolic pathways may offer promising avenues for the prevention and management of Childhood with obesity. Future research should further elucidate the precise biological mechanisms by which these gut microbiome-derived metabolites influence obesity risk. Additionally, exploring the potential for personalized interventions that modulate the gut microbiome and its metabolic outputs could lead to more effective strategies for tackling the Childhood with obesity epidemic. In conclusion, this study has uncovered compelling causal evidence linking gut microbiome-associated blood metabolites to the risk of Childhood with obesity. These insights pave the way for developing novel, microbiome-based approaches to address this pressing public health challenge.

The present study has several strengths that can guide future directions in Childhood with obesity research. First, our work comprises the first systematic investigation to examine the causal relationship between gut microbiota, blood metabolites, and Childhood with obesity. This unique contribution is underscored by the comprehensive analysis of 473 distinct types of gut microbiota and 233 blood metabolites. By using an MR design, the study effectively mitigated issues related to reverse causality and residual confounding variables.^48^ Furthermore, thorough sensitivity analyses were conducted to eliminate the potential influence of genetic polymorphisms, thereby enhancing the validity of the causal inferences drawn from the study. Second, compared with conventional ideas of studying gut microbiota and metabolites within the Childhood with obesity microenvironment, our study aimed to examine the causal relationship between gut microbiota, blood metabolites and Childhood with obesity. Our findings underscore the potential impact of gut microbiota and metabolites in peripheral blood on the progression of Childhood with obesity, shedding light on their role in the onset and advancement of the disease. Third, we have identified a set of gut microbiota and blood metabolites that exhibit a strong correlation with the risk of Childhood with obesity. These findings present potential biomarkers for non-invasive stool testing in Childhood with obesity that warrant validation through subsequent experiments. Finally, we have proposed a potential axis of K10 sp001941205–Total cholesterol to total lipids ratio in medium VLDL–Childhood with obesity, as well as another axis SM23-33–Ratio of 22:6 docosahexaenoic acid to total fatty acids–Childhood with obesity, which may serve as a foundation for drug development strategies in Childhood with obesity research. Further investigation utilizing in vitro and in vivo models is necessary to validate these proposed axes and to develop targeted therapies accordingly.

The present study performed a two-sample, bidirectional, multivariable, and two-step mediation MR analysis utilizing large genome-wide association study datasets, demonstrating high statistical efficiency. The conclusions drawn in our study are grounded in genetic IVs, with causal inference conducted through multiple MR analysis methods. The findings are robust and unaffected by horizontal pleiotropy and other confounding factors. Meanwhile, our study also has several limitations. First, our analysis was performed using the European population, which limits its application in all people worldwide. Second, the dataset on Childhood with obesity is general and lacks subtype information, which may not accurately reflect the characteristics of specific subtypes within Childhood with obesity. Third, despite efforts to detect and remove outlier variants, the potential influence of horizontal pleiotropy on our findings cannot be completely ruled out. Additionally, the utilization of summary-level statistics rather than individual-level data in our analysis limits our ability to investigate causal relationships within specific subgroups, such as females and males. Finally, our research indicates a modest genetic prediction of Childhood with obesity, mediated by the total cholesterol to total lipids ratio in medium VLDL at 2.53% (95%CI; 2.14%-2.92%) and the ratio of 22:6 docosahexaenoic acid to total fatty acids at 4.07% (95%CI; 2.70%-5.44%), suggesting the need for further investigation into additional mediators.

## Data Availability

The analysis utilized publicly available datasets. Detailed information on all original contributions can be found in the "Data Sources" section, including specific download links and accession numbers. Readers can refer to this section for access. For further inquiries, please contact the corresponding author.

https://ftp.ebi.ac.uk/pub/databases/gwas/summary_statistics/

http://egg-consortium.org/childhood-obesity-2019.html

## Authors Contributions

Min Zhang conceived and designed the study, conducted data analysis and interpretation, and wrote the manuscript. Wenjuan Yan developed the research hypothesis.

