## Supplementary Tables S1-S8 for "Genetically Predicted Blood Metabolites Mediate the Association Between Gut Microbiota and Childhood with obesity: A Mendelian Randomization Study"

| **Table S1. STROBE-MR checklist of recommended items to address in reports of Mendelian randomization studies^1-2^** | | |
| --- | --- | --- |
| **Item No.** | **Section** | **Checklist item** |
| 1 | **TITLE and ABSTRACT** | Indicate Mendelian randomization (MR) as the study’s design in the title and/or the abstract if that is a main purpose of the study |
|  | **INTRODUCTION** | |
| 2 | **Background** | Explain the scientific background and rationale for the reported study. What is the exposure? Is a potential causal relationship between exposure and outcome plausible? Justify why MR is a helpful method to address the study question |
| 3 | **Objectives** | State specific objectives clearly, including pre-specified causal hypotheses (if any). State that MR is a method that, under specific assumptions, intends to estimate causal effects |
|  | **METHODS** | |
| 4 | **Study design and data sources** | Present key elements of the study design early in the article. Consider including a table listing sources of data for all phases of the study. For each data source contributing to the analysis, describe the following: |
|  | a) | Setting: Describe the study design and the underlying population, if possible. Describe the setting, locations, and relevant dates, including periods of recruitment, exposure, follow-up, and data collection, when available. |
|  | b) | Participants: Give the eligibility criteria, and the sources and methods of selection of participants. Report the sample size, and whether any power or sample size calculations were carried out prior to the main analysis |
|  | c) | Describe measurement, quality control and selection of genetic variants |
|  | d) | For each exposure, outcome, and other relevant variables, describe methods of assessment and diagnostic criteria for diseases |
|  | e) | Provide details of ethics committee approval and participant informed consent, if relevant |
| 5 | **Assumptions** | Explicitly state the three core IV assumptions for the main analysis (relevance, independence and exclusion restriction) as well assumptions for any additional or sensitivity analysis |
| 6 | **Statistical methods: main analysis** | Describe statistical methods and statistics used |
|  | a) | Describe how quantitative variables were handled in the analyses (i.e., scale, units, model) |
|  | b) | Describe how genetic variants were handled in the analyses and, if applicable, how their weights were selected |
|  | c) | Describe the MR estimator (e.g. two-stage least squares, Wald ratio) and related statistics. Detail the included covariates and, in case of two-sample MR, whether the same covariate set was used for adjustment in the two samples |
|  | d) | Explain how missing data were addressed |
|  | e) | If applicable, indicate how multiple testing was addressed |
| 7 | **Assessment of assumptions** | Describe any methods or prior knowledge used to assess the assumptions or justify their validity |
| 8 | **Sensitivity analyses and additional analyses** | Describe any sensitivity analyses or additional analyses performed (e.g. comparison of effect estimates from different approaches, independent replication, bias analytic techniques, validation of instruments, simulations) |
| 9 | **Software and pre-registration** | |
|  | a) | Name statistical software and package(s), including version and settings used |
|  | b) | State whether the study protocol and details were pre-registered (as well as when and where) |
|  | **RESULTS** | |
| 10 | **Descriptive data** | |
|  | a) | Report the numbers of individuals at each stage of included studies and reasons for exclusion. Consider use of a flow diagram |
|  | b) | Report summary statistics for phenotypic exposure(s), outcome(s), and other relevant variables (e.g. means, SDs, proportions) |
|  | c) | If the data sources include meta-analyses of previous studies, provide the assessments of heterogeneity across these studies |
|  | d) | For two-sample MR: |
|  |  | i. Provide justification of the similarity of the genetic variant-exposure associations between the exposure and outcome samples |
|  |  | ii.  Provide information on the number of individuals who overlap between the exposure and outcome studies |
| 11 | **Main results** | |
|  | a) | Report the associations between genetic variant and exposure, and between genetic variant and outcome, preferably on an interpretable scale |
|  | b) | Report MR estimates of the relationship between exposure and outcome, and the measures of uncertainty from the MR analysis, on an interpretable scale, such as odds ratio or relative risk per SD difference |
|  | c) | If relevant, consider translating estimates of relative risk into absolute risk for a meaningful time period |
|  | d) | Consider plots to visualize results (e.g. forest plot, scatterplot of associations between genetic variants and outcome versus between genetic variants and exposure) |
| 12 | **Assessment of assumptions** | |
|  | a) | Report the assessment of the validity of the assumptions |
|  | b) | Report any additional statistics (e.g., assessments of heterogeneity across genetic variants, such as I2, Q statistic or E-value) |
| 13 | **Sensitivity analyses and additional analyses** | |
|  | a) | Report any sensitivity analyses to assess the robustness of the main results to violations of the assumptions |
|  | b) | Report results from other sensitivity analyses or additional analyses |
|  | c) | Report any assessment of direction of causal relationship (e.g., bidirectional MR) |
|  | d) | When relevant, report and compare with estimates from non-MR analyses |
|  | e) | Consider additional plots to visualize results (e.g., leave-one-out analyses) |
|  | **DISCUSSION** | |
| 14 | **Key results** | Summarize key results with reference to study objectives |
| 15 | **Limitations** | Discuss limitations of the study, taking into account the validity of the IV assumptions, other sources of potential bias, and imprecision. Discuss both direction and magnitude of any potential bias and any efforts to address them |
| 16 | **Interpretation** | |
|  | a) | Meaning: Give a cautious overall interpretation of results in the context of their limitations and in comparison with other studies |
|  | b) | Mechanism: Discuss underlying biological mechanisms that could drive a potential causal relationship between the investigated exposure and the outcome, and whether the gene-environment equivalence assumption is reasonable. Use causal language carefully, clarifying that IV estimates may provide causal effects only under certain assumptions |
|  | c) | Clinical relevance: Discuss whether the results have clinical or public policy relevance, and to what extent they inform effect sizes of possible interventions |
| 17 | **Generalizability** | Discuss the generalizability of the study results (a) to other populations, (b) across other exposure periods/timings, and (c) across other levels of exposure |
|  | **OTHER INFORMATION** | |
| 18 | **Funding** | Describe sources of funding and the role of funders in the present study and, if applicable, sources of funding for the databases and original study or studies on which the present study is based |
| 19 | **Data and data sharing** | Provide the data used to perform all analyses or report where and how the data can be accessed, and reference these sources in the article. Provide the statistical code needed to reproduce the results in the article, or report whether the code is publicly accessible and if so, where |
| 20 | **Conflicts of Interest** | All authors should declare all potential conflicts of interest |
| This checklist is copyrighted by the Equator Network under the Creative Commons Attribution 3.0 Unported (CC BY 3.0) license. 1. Skrivankova VW, Richmond RC, Woolf BAR, Yarmolinsky J, Davies NM, Swanson SA, et al. Strengthening the Reporting of Observational Studies in Epidemiology using Mendelian Randomization (STROBE-MR) Statement. JAMA. 2021;under review.  2. Skrivankova VW, Richmond RC, Woolf BAR, Davies NM, Swanson SA, VanderWeele TJ, et al. Strengthening the Reporting of Observational Studies in Epidemiology using Mendelian Randomisation (STROBE-MR): Explanation and Elaboration. BMJ. 2021;375:n2233. | | |

| **Table S2. Results of Mendelian Randomization Analysis of Gut Microbiota on Childhood Obesity** | | | | | | | | | | | |
| --- | --- | --- | --- | --- | --- | --- | --- | --- | --- | --- | --- |
| **exposure** | **outcome** | **method** | **nsnp** | **b** | **se** | **pval** | **lo_ci** | **up_ci** | **or** | **or_lci95** | **or_uci95** |
| An181 | Childhood Obesity | Inverse variance weighted | 70 | 0.140 | 0.065 | 0.030 | 0.013 | 0.267 | 1.151 | 1.013 | 1.307 |
| An181 | Childhood Obesity | MR Egger | 70 | 0.289 | 0.164 | 0.083 | -0.033 | 0.611 | 1.335 | 0.968 | 1.842 |
| An181 | Childhood Obesity | Simple mode | 70 | 0.195 | 0.220 | 0.380 | -0.237 | 0.627 | 1.215 | 0.789 | 1.871 |
| An181 | Childhood Obesity | Weighted median | 70 | 0.179 | 0.089 | 0.044 | 0.005 | 0.354 | 1.197 | 1.005 | 1.424 |
| An181 | Childhood Obesity | Weighted mode | 70 | 0.210 | 0.200 | 0.296 | -0.181 | 0.601 | 1.234 | 0.834 | 1.825 |
| Bacteroides A | Childhood Obesity | Inverse variance weighted | 56 | -0.070 | 0.033 | 0.033 | -0.135 | -0.006 | 0.932 | 0.873 | 0.994 |
| Bacteroides A | Childhood Obesity | MR Egger | 56 | -0.120 | 0.088 | 0.175 | -0.292 | 0.051 | 0.886 | 0.747 | 1.052 |
| Bacteroides A | Childhood Obesity | Simple mode | 56 | -0.070 | 0.094 | 0.460 | -0.255 | 0.115 | 0.932 | 0.775 | 1.122 |
| Bacteroides A | Childhood Obesity | Weighted median | 56 | -0.063 | 0.047 | 0.179 | -0.156 | 0.029 | 0.939 | 0.856 | 1.029 |
| Bacteroides A | Childhood Obesity | Weighted mode | 56 | -0.063 | 0.090 | 0.490 | -0.239 | 0.114 | 0.939 | 0.787 | 1.121 |
| CAG-698 | Childhood Obesity | Inverse variance weighted | 70 | 0.223 | 0.070 | 0.001 | 0.087 | 0.360 | 1.250 | 1.091 | 1.433 |
| CAG-698 | Childhood Obesity | MR Egger | 70 | 0.214 | 0.146 | 0.146 | -0.071 | 0.500 | 1.239 | 0.931 | 1.649 |
| CAG-698 | Childhood Obesity | Simple mode | 70 | 0.288 | 0.232 | 0.219 | -0.167 | 0.743 | 1.333 | 0.846 | 2.101 |
| CAG-698 | Childhood Obesity | Weighted median | 70 | 0.175 | 0.093 | 0.058 | -0.006 | 0.356 | 1.191 | 0.994 | 1.428 |
| CAG-698 | Childhood Obesity | Weighted mode | 70 | 0.297 | 0.191 | 0.124 | -0.077 | 0.671 | 1.346 | 0.926 | 1.957 |
| Coprobacillus | Childhood Obesity | Inverse variance weighted | 63 | -0.105 | 0.047 | 0.025 | -0.196 | -0.013 | 0.901 | 0.822 | 0.987 |
| Coprobacillus | Childhood Obesity | MR Egger | 63 | -0.009 | 0.101 | 0.925 | -0.207 | 0.189 | 0.991 | 0.813 | 1.207 |
| Coprobacillus | Childhood Obesity | Simple mode | 63 | -0.072 | 0.156 | 0.648 | -0.377 | 0.234 | 0.931 | 0.686 | 1.264 |
| Coprobacillus | Childhood Obesity | Weighted median | 63 | -0.089 | 0.067 | 0.185 | -0.220 | 0.043 | 0.915 | 0.802 | 1.043 |
| Coprobacillus | Childhood Obesity | Weighted mode | 63 | -0.079 | 0.174 | 0.651 | -0.421 | 0.263 | 0.924 | 0.656 | 1.300 |
| Eubacterium I ramulus A | Childhood Obesity | Inverse variance weighted | 65 | -0.168 | 0.066 | 0.011 | -0.297 | -0.038 | 0.846 | 0.743 | 0.963 |
| Eubacterium I ramulus A | Childhood Obesity | MR Egger | 65 | -0.098 | 0.177 | 0.582 | -0.446 | 0.250 | 0.906 | 0.640 | 1.283 |
| Eubacterium I ramulus A | Childhood Obesity | Simple mode | 65 | -0.355 | 0.234 | 0.134 | -0.813 | 0.104 | 0.701 | 0.444 | 1.109 |
| Eubacterium I ramulus A | Childhood Obesity | Weighted median | 65 | -0.160 | 0.093 | 0.086 | -0.343 | 0.023 | 0.852 | 0.709 | 1.023 |
| Eubacterium I ramulus A | Childhood Obesity | Weighted mode | 65 | -0.155 | 0.230 | 0.503 | -0.605 | 0.296 | 0.857 | 0.546 | 1.344 |
| Faecalicatena sp001517425 | Childhood Obesity | Inverse variance weighted | 60 | 0.217 | 0.084 | 0.010 | 0.051 | 0.382 | 1.242 | 1.053 | 1.465 |
| Faecalicatena sp001517425 | Childhood Obesity | MR Egger | 60 | 0.226 | 0.182 | 0.218 | -0.130 | 0.582 | 1.254 | 0.878 | 1.790 |
| Faecalicatena sp001517425 | Childhood Obesity | Simple mode | 60 | 0.224 | 0.274 | 0.417 | -0.313 | 0.761 | 1.251 | 0.731 | 2.141 |
| Faecalicatena sp001517425 | Childhood Obesity | Weighted median | 60 | 0.164 | 0.162 | 0.312 | -0.154 | 0.482 | 1.178 | 0.857 | 1.620 |
| Faecalicatena sp001517425 | Childhood Obesity | Weighted mode | 60 | 0.182 | 0.167 | 0.279 | -0.144 | 0.509 | 1.200 | 0.866 | 1.663 |
| Halomonadaceae | Childhood Obesity | Inverse variance weighted | 79 | 0.405 | 0.204 | 0.047 | 0.005 | 0.805 | 1.499 | 1.005 | 2.236 |
| Halomonadaceae | Childhood Obesity | MR Egger | 79 | 0.833 | 0.547 | 0.132 | -0.239 | 1.904 | 2.299 | 0.787 | 6.716 |
| Halomonadaceae | Childhood Obesity | Simple mode | 79 | 0.077 | 0.749 | 0.918 | -1.391 | 1.545 | 1.080 | 0.249 | 4.690 |
| Halomonadaceae | Childhood Obesity | Weighted median | 79 | 0.383 | 0.294 | 0.192 | -0.192 | 0.959 | 1.467 | 0.825 | 2.609 |
| Halomonadaceae | Childhood Obesity | Weighted mode | 79 | 0.111 | 0.677 | 0.870 | -1.216 | 1.439 | 1.118 | 0.296 | 4.215 |
| K10 sp001941205 | Childhood Obesity | Inverse variance weighted | 68 | -0.110 | 0.055 | 0.043 | -0.217 | -0.003 | 0.895 | 0.805 | 0.997 |
| K10 sp001941205 | Childhood Obesity | MR Egger | 68 | -0.110 | 0.129 | 0.400 | -0.363 | 0.144 | 0.896 | 0.695 | 1.155 |
| K10 sp001941205 | Childhood Obesity | Simple mode | 68 | -0.041 | 0.175 | 0.816 | -0.383 | 0.302 | 0.960 | 0.682 | 1.352 |
| K10 sp001941205 | Childhood Obesity | Weighted median | 68 | -0.087 | 0.074 | 0.239 | -0.232 | 0.058 | 0.916 | 0.793 | 1.060 |
| K10 sp001941205 | Childhood Obesity | Weighted mode | 68 | -0.080 | 0.145 | 0.581 | -0.364 | 0.203 | 0.923 | 0.695 | 1.225 |
| Ruminococcus E sp900314705 | Childhood Obesity | Inverse variance weighted | 87 | -0.091 | 0.045 | 0.046 | -0.180 | -0.002 | 0.913 | 0.836 | 0.998 |
| Ruminococcus E sp900314705 | Childhood Obesity | MR Egger | 87 | -0.148 | 0.097 | 0.129 | -0.338 | 0.041 | 0.862 | 0.713 | 1.042 |
| Ruminococcus E sp900314705 | Childhood Obesity | Simple mode | 87 | -0.218 | 0.179 | 0.225 | -0.569 | 0.132 | 0.804 | 0.566 | 1.141 |
| Ruminococcus E sp900314705 | Childhood Obesity | Weighted median | 87 | -0.133 | 0.066 | 0.043 | -0.261 | -0.004 | 0.876 | 0.770 | 0.996 |
| Ruminococcus E sp900314705 | Childhood Obesity | Weighted mode | 87 | -0.204 | 0.157 | 0.196 | -0.512 | 0.103 | 0.815 | 0.599 | 1.109 |
| SM23-33 | Childhood Obesity | Inverse variance weighted | 71 | -0.282 | 0.118 | 0.017 | -0.513 | -0.050 | 0.755 | 0.599 | 0.951 |
| SM23-33 | Childhood Obesity | MR Egger | 71 | -0.261 | 0.304 | 0.393 | -0.857 | 0.335 | 0.770 | 0.425 | 1.397 |
| SM23-33 | Childhood Obesity | Simple mode | 71 | -0.080 | 0.387 | 0.837 | -0.839 | 0.679 | 0.923 | 0.432 | 1.972 |
| SM23-33 | Childhood Obesity | Weighted median | 71 | -0.142 | 0.169 | 0.400 | -0.473 | 0.189 | 0.868 | 0.623 | 1.208 |
| SM23-33 | Childhood Obesity | Weighted mode | 71 | -0.080 | 0.342 | 0.816 | -0.751 | 0.591 | 0.923 | 0.472 | 1.806 |
| TMED109 | Childhood Obesity | Inverse variance weighted | 64 | -0.232 | 0.098 | 0.018 | -0.424 | -0.040 | 0.793 | 0.654 | 0.961 |
| TMED109 | Childhood Obesity | MR Egger | 64 | -0.473 | 0.236 | 0.049 | -0.935 | -0.011 | 0.623 | 0.393 | 0.989 |
| TMED109 | Childhood Obesity | Simple mode | 64 | -0.453 | 0.319 | 0.160 | -1.077 | 0.172 | 0.636 | 0.341 | 1.187 |
| TMED109 | Childhood Obesity | Weighted median | 64 | -0.362 | 0.140 | 0.010 | -0.637 | -0.088 | 0.696 | 0.529 | 0.915 |
| TMED109 | Childhood Obesity | Weighted mode | 64 | -0.441 | 0.292 | 0.135 | -1.013 | 0.130 | 0.643 | 0.363 | 1.139 |
| UBA1206 sp000433115 | Childhood Obesity | Inverse variance weighted | 68 | -0.077 | 0.038 | 0.043 | -0.152 | -0.002 | 0.926 | 0.859 | 0.998 |
| UBA1206 sp000433115 | Childhood Obesity | MR Egger | 68 | -0.075 | 0.084 | 0.378 | -0.241 | 0.091 | 0.928 | 0.786 | 1.095 |
| UBA1206 sp000433115 | Childhood Obesity | Simple mode | 68 | -0.108 | 0.124 | 0.387 | -0.351 | 0.135 | 0.898 | 0.704 | 1.145 |
| UBA1206 sp000433115 | Childhood Obesity | Weighted median | 68 | -0.066 | 0.055 | 0.227 | -0.174 | 0.041 | 0.936 | 0.841 | 1.042 |
| UBA1206 sp000433115 | Childhood Obesity | Weighted mode | 68 | -0.057 | 0.132 | 0.666 | -0.315 | 0.201 | 0.944 | 0.729 | 1.223 |
| UBA8621 | Childhood Obesity | Inverse variance weighted | 66 | 0.226 | 0.104 | 0.030 | 0.022 | 0.430 | 1.254 | 1.022 | 1.538 |
| UBA8621 | Childhood Obesity | MR Egger | 66 | 0.037 | 0.262 | 0.887 | -0.476 | 0.551 | 1.038 | 0.621 | 1.735 |
| UBA8621 | Childhood Obesity | Simple mode | 66 | 0.224 | 0.336 | 0.507 | -0.435 | 0.883 | 1.252 | 0.647 | 2.419 |
| UBA8621 | Childhood Obesity | Weighted median | 66 | 0.222 | 0.150 | 0.139 | -0.072 | 0.515 | 1.248 | 0.931 | 1.674 |
| UBA8621 | Childhood Obesity | Weighted mode | 66 | 0.201 | 0.336 | 0.553 | -0.459 | 0.860 | 1.222 | 0.632 | 2.363 |

| **Table S3. Results of Mendelian Randomization Analysis of Childhood Obesity on Gut Microbiota** | | | | | | | | | | | |
| --- | --- | --- | --- | --- | --- | --- | --- | --- | --- | --- | --- |
| **exposure** | **outcome** | **method** | **nsnp** | **b** | **se** | **pval** | **lo_ci** | **up_ci** | **or** | **or_lci95** | **or_uci95** |
| Childhood Obesity | An181 | Inverse variance weighted | 64 | -0.009 | 0.011 | 0.402 | -0.030 | 0.012 | 0.991 | 0.970 | 1.012 |
| Childhood Obesity | An181 | MR Egger | 64 | -0.022 | 0.027 | 0.433 | -0.075 | 0.032 | 0.979 | 0.927 | 1.033 |
| Childhood Obesity | An181 | Simple mode | 64 | 0.000 | 0.038 | 0.994 | -0.075 | 0.075 | 1.000 | 0.927 | 1.078 |
| Childhood Obesity | An181 | Weighted median | 64 | -0.002 | 0.016 | 0.888 | -0.034 | 0.029 | 0.998 | 0.967 | 1.030 |
| Childhood Obesity | An181 | Weighted mode | 64 | 0.002 | 0.032 | 0.952 | -0.061 | 0.065 | 1.002 | 0.940 | 1.068 |
| Childhood Obesity | Bacteroides A | Inverse variance weighted | 64 | 0.010 | 0.026 | 0.690 | -0.040 | 0.061 | 1.010 | 0.960 | 1.063 |
| Childhood Obesity | Bacteroides A | MR Egger | 64 | 0.097 | 0.066 | 0.145 | -0.032 | 0.226 | 1.102 | 0.969 | 1.253 |
| Childhood Obesity | Bacteroides A | Simple mode | 64 | -0.036 | 0.080 | 0.658 | -0.192 | 0.121 | 0.965 | 0.825 | 1.129 |
| Childhood Obesity | Bacteroides A | Weighted median | 64 | -0.003 | 0.036 | 0.933 | -0.073 | 0.067 | 0.997 | 0.930 | 1.069 |
| Childhood Obesity | Bacteroides A | Weighted mode | 64 | -0.005 | 0.073 | 0.948 | -0.147 | 0.138 | 0.995 | 0.863 | 1.147 |
| Childhood Obesity | CAG-698 | Inverse variance weighted | 64 | -0.005 | 0.012 | 0.661 | -0.029 | 0.018 | 0.995 | 0.972 | 1.018 |
| Childhood Obesity | CAG-698 | MR Egger | 64 | -0.018 | 0.031 | 0.572 | -0.078 | 0.043 | 0.983 | 0.925 | 1.044 |
| Childhood Obesity | CAG-698 | Simple mode | 64 | 0.001 | 0.043 | 0.984 | -0.083 | 0.085 | 1.001 | 0.920 | 1.089 |
| Childhood Obesity | CAG-698 | Weighted median | 64 | -0.013 | 0.017 | 0.447 | -0.046 | 0.020 | 0.987 | 0.955 | 1.021 |
| Childhood Obesity | CAG-698 | Weighted mode | 64 | -0.011 | 0.037 | 0.765 | -0.083 | 0.061 | 0.989 | 0.921 | 1.062 |
| Childhood Obesity | Coprobacillus | Inverse variance weighted | 64 | 0.006 | 0.015 | 0.707 | -0.024 | 0.035 | 1.006 | 0.976 | 1.036 |
| Childhood Obesity | Coprobacillus | MR Egger | 64 | -0.027 | 0.038 | 0.484 | -0.103 | 0.048 | 0.973 | 0.903 | 1.050 |
| Childhood Obesity | Coprobacillus | Simple mode | 64 | 0.031 | 0.046 | 0.504 | -0.060 | 0.122 | 1.032 | 0.942 | 1.130 |
| Childhood Obesity | Coprobacillus | Weighted median | 64 | 0.018 | 0.021 | 0.377 | -0.023 | 0.059 | 1.019 | 0.978 | 1.061 |
| Childhood Obesity | Coprobacillus | Weighted mode | 64 | 0.027 | 0.036 | 0.456 | -0.044 | 0.098 | 1.027 | 0.957 | 1.103 |
| Childhood Obesity | Eubacterium I ramulus A | Inverse variance weighted | 64 | 0.002 | 0.011 | 0.845 | -0.019 | 0.024 | 1.002 | 0.981 | 1.024 |
| Childhood Obesity | Eubacterium I ramulus A | MR Egger | 64 | 0.024 | 0.028 | 0.395 | -0.031 | 0.079 | 1.024 | 0.969 | 1.082 |
| Childhood Obesity | Eubacterium I ramulus A | Simple mode | 64 | -0.036 | 0.034 | 0.299 | -0.103 | 0.031 | 0.965 | 0.902 | 1.032 |
| Childhood Obesity | Eubacterium I ramulus A | Weighted median | 64 | -0.017 | 0.016 | 0.285 | -0.049 | 0.014 | 0.983 | 0.952 | 1.014 |
| Childhood Obesity | Eubacterium I ramulus A | Weighted mode | 64 | -0.026 | 0.029 | 0.373 | -0.083 | 0.031 | 0.974 | 0.921 | 1.031 |
| Childhood Obesity | Faecalicatena sp001517425 | Inverse variance weighted | 64 | 0.005 | 0.008 | 0.486 | -0.010 | 0.020 | 1.005 | 0.990 | 1.021 |
| Childhood Obesity | Faecalicatena sp001517425 | MR Egger | 64 | 0.000 | 0.020 | 0.982 | -0.039 | 0.038 | 1.000 | 0.962 | 1.039 |
| Childhood Obesity | Faecalicatena sp001517425 | Simple mode | 64 | -0.008 | 0.023 | 0.747 | -0.053 | 0.038 | 0.993 | 0.948 | 1.039 |
| Childhood Obesity | Faecalicatena sp001517425 | Weighted median | 64 | 0.003 | 0.011 | 0.799 | -0.018 | 0.024 | 1.003 | 0.982 | 1.024 |
| Childhood Obesity | Faecalicatena sp001517425 | Weighted mode | 64 | -0.002 | 0.019 | 0.925 | -0.040 | 0.036 | 0.998 | 0.961 | 1.037 |
| Childhood Obesity | Halomonadaceae | Inverse variance weighted | 64 | -0.006 | 0.004 | 0.105 | -0.013 | 0.001 | 0.994 | 0.987 | 1.001 |
| Childhood Obesity | Halomonadaceae | MR Egger | 64 | -0.012 | 0.009 | 0.194 | -0.030 | 0.006 | 0.988 | 0.971 | 1.006 |
| Childhood Obesity | Halomonadaceae | Simple mode | 64 | -0.008 | 0.013 | 0.551 | -0.032 | 0.017 | 0.992 | 0.968 | 1.017 |
| Childhood Obesity | Halomonadaceae | Weighted median | 64 | -0.006 | 0.005 | 0.287 | -0.016 | 0.005 | 0.994 | 0.984 | 1.005 |
| Childhood Obesity | Halomonadaceae | Weighted mode | 64 | -0.007 | 0.011 | 0.503 | -0.028 | 0.014 | 0.993 | 0.972 | 1.014 |
| Childhood Obesity | K10 sp001941205 | Inverse variance weighted | 64 | 0.023 | 0.016 | 0.152 | -0.008 | 0.054 | 1.023 | 0.992 | 1.056 |
| Childhood Obesity | K10 sp001941205 | MR Egger | 64 | 0.004 | 0.041 | 0.927 | -0.077 | 0.084 | 1.004 | 0.926 | 1.088 |
| Childhood Obesity | K10 sp001941205 | Simple mode | 64 | 0.044 | 0.048 | 0.366 | -0.050 | 0.138 | 1.045 | 0.951 | 1.148 |
| Childhood Obesity | K10 sp001941205 | Weighted median | 64 | 0.043 | 0.021 | 0.039 | 0.002 | 0.084 | 1.044 | 1.002 | 1.087 |
| Childhood Obesity | K10 sp001941205 | Weighted mode | 64 | 0.057 | 0.038 | 0.135 | -0.017 | 0.132 | 1.059 | 0.983 | 1.141 |
| Childhood Obesity | Ruminococcus E sp900314705 | Inverse variance weighted | 64 | 0.022 | 0.014 | 0.104 | -0.005 | 0.050 | 1.023 | 0.995 | 1.051 |
| Childhood Obesity | Ruminococcus E sp900314705 | MR Egger | 64 | 0.037 | 0.035 | 0.304 | -0.033 | 0.106 | 1.037 | 0.968 | 1.112 |
| Childhood Obesity | Ruminococcus E sp900314705 | Simple mode | 64 | 0.045 | 0.044 | 0.312 | -0.041 | 0.131 | 1.046 | 0.959 | 1.140 |
| Childhood Obesity | Ruminococcus E sp900314705 | Weighted median | 64 | 0.036 | 0.020 | 0.074 | -0.003 | 0.075 | 1.036 | 0.997 | 1.078 |
| Childhood Obesity | Ruminococcus E sp900314705 | Weighted mode | 64 | 0.045 | 0.038 | 0.248 | -0.030 | 0.120 | 1.046 | 0.970 | 1.128 |
| Childhood Obesity | SM23-33 | Inverse variance weighted | 64 | -0.012 | 0.007 | 0.069 | -0.025 | 0.001 | 0.988 | 0.975 | 1.001 |
| Childhood Obesity | SM23-33 | MR Egger | 64 | -0.028 | 0.017 | 0.104 | -0.061 | 0.005 | 0.972 | 0.941 | 1.005 |
| Childhood Obesity | SM23-33 | Simple mode | 64 | -0.008 | 0.023 | 0.715 | -0.053 | 0.036 | 0.992 | 0.949 | 1.037 |
| Childhood Obesity | SM23-33 | Weighted median | 64 | -0.008 | 0.009 | 0.383 | -0.026 | 0.010 | 0.992 | 0.975 | 1.010 |
| Childhood Obesity | SM23-33 | Weighted mode | 64 | -0.002 | 0.017 | 0.893 | -0.035 | 0.030 | 0.998 | 0.966 | 1.031 |
| Childhood Obesity | TMED109 | Inverse variance weighted | 64 | 0.005 | 0.008 | 0.543 | -0.010 | 0.019 | 1.005 | 0.990 | 1.019 |
| Childhood Obesity | TMED109 | MR Egger | 64 | 0.024 | 0.019 | 0.217 | -0.014 | 0.062 | 1.024 | 0.986 | 1.063 |
| Childhood Obesity | TMED109 | Simple mode | 64 | 0.009 | 0.028 | 0.746 | -0.045 | 0.064 | 1.009 | 0.956 | 1.066 |
| Childhood Obesity | TMED109 | Weighted median | 64 | 0.005 | 0.011 | 0.624 | -0.016 | 0.027 | 1.005 | 0.984 | 1.027 |
| Childhood Obesity | TMED109 | Weighted mode | 64 | 0.019 | 0.023 | 0.411 | -0.027 | 0.065 | 1.020 | 0.974 | 1.067 |
| Childhood Obesity | UBA1206 sp000433115 | Inverse variance weighted | 64 | -0.007 | 0.018 | 0.712 | -0.043 | 0.029 | 0.993 | 0.958 | 1.030 |
| Childhood Obesity | UBA1206 sp000433115 | MR Egger | 64 | -0.004 | 0.047 | 0.933 | -0.096 | 0.088 | 0.996 | 0.908 | 1.092 |
| Childhood Obesity | UBA1206 sp000433115 | Simple mode | 64 | -0.019 | 0.055 | 0.732 | -0.127 | 0.089 | 0.981 | 0.880 | 1.094 |
| Childhood Obesity | UBA1206 sp000433115 | Weighted median | 64 | -0.006 | 0.026 | 0.806 | -0.058 | 0.045 | 0.994 | 0.944 | 1.046 |
| Childhood Obesity | UBA1206 sp000433115 | Weighted mode | 64 | -0.017 | 0.049 | 0.728 | -0.113 | 0.079 | 0.983 | 0.893 | 1.082 |
| Childhood Obesity | UBA8621 | Inverse variance weighted | 64 | 0.003 | 0.007 | 0.661 | -0.010 | 0.016 | 1.003 | 0.990 | 1.016 |
| Childhood Obesity | UBA8621 | MR Egger | 64 | 0.003 | 0.017 | 0.881 | -0.031 | 0.036 | 1.003 | 0.970 | 1.037 |
| Childhood Obesity | UBA8621 | Simple mode | 64 | -0.003 | 0.021 | 0.881 | -0.045 | 0.039 | 0.997 | 0.956 | 1.040 |
| Childhood Obesity | UBA8621 | Weighted median | 64 | 0.001 | 0.010 | 0.900 | -0.018 | 0.021 | 1.001 | 0.982 | 1.021 |
| Childhood Obesity | UBA8621 | Weighted mode | 64 | 0.001 | 0.018 | 0.959 | -0.035 | 0.036 | 1.001 | 0.966 | 1.037 |

| **Table S4. Results of Mendelian Randomization Analysis of Blood Metabolites on Childhood Obesity** | | | | | | | | | | | |
| --- | --- | --- | --- | --- | --- | --- | --- | --- | --- | --- | --- |
| **exposure** | **outcome** | **method** | **nsnp** | **b** | **se** | **pval** | **lo_ci** | **up_ci** | **or** | **or_lci95** | **or_uci95** |
| Albumin levels | Childhood Obesity | Inverse variance weighted | 18 | -0.332 | 0.160 | 0.037 | -0.646 | -0.019 | 0.717 | 0.524 | 0.981 |
| Albumin levels | Childhood Obesity | MR Egger | 18 | -0.171 | 0.353 | 0.635 | -0.862 | 0.521 | 0.843 | 0.422 | 1.683 |
| Albumin levels | Childhood Obesity | Simple mode | 18 | -0.525 | 0.405 | 0.212 | -1.319 | 0.269 | 0.592 | 0.267 | 1.309 |
| Albumin levels | Childhood Obesity | Weighted median | 18 | -0.275 | 0.225 | 0.222 | -0.716 | 0.166 | 0.760 | 0.489 | 1.180 |
| Albumin levels | Childhood Obesity | Weighted mode | 18 | -0.395 | 0.355 | 0.281 | -1.091 | 0.301 | 0.674 | 0.336 | 1.351 |
| Cholesteryl esters to total lipids ratio in small HDL | Childhood Obesity | Inverse variance weighted | 64 | -0.147 | 0.055 | 0.008 | -0.256 | -0.039 | 0.863 | 0.774 | 0.962 |
| Cholesteryl esters to total lipids ratio in small HDL | Childhood Obesity | MR Egger | 64 | -0.108 | 0.084 | 0.205 | -0.272 | 0.057 | 0.898 | 0.762 | 1.059 |
| Cholesteryl esters to total lipids ratio in small HDL | Childhood Obesity | Simple mode | 64 | -0.256 | 0.174 | 0.147 | -0.596 | 0.085 | 0.774 | 0.551 | 1.089 |
| Cholesteryl esters to total lipids ratio in small HDL | Childhood Obesity | Weighted median | 64 | -0.205 | 0.086 | 0.017 | -0.372 | -0.037 | 0.815 | 0.689 | 0.964 |
| Cholesteryl esters to total lipids ratio in small HDL | Childhood Obesity | Weighted mode | 64 | -0.198 | 0.092 | 0.035 | -0.378 | -0.017 | 0.821 | 0.685 | 0.983 |
| Cholesteryl esters to total lipids ratio in small LDL | Childhood Obesity | Inverse variance weighted | 65 | -0.103 | 0.051 | 0.042 | -0.203 | -0.004 | 0.902 | 0.817 | 0.996 |
| Cholesteryl esters to total lipids ratio in small LDL | Childhood Obesity | MR Egger | 65 | -0.108 | 0.080 | 0.182 | -0.265 | 0.049 | 0.897 | 0.767 | 1.050 |
| Cholesteryl esters to total lipids ratio in small LDL | Childhood Obesity | Simple mode | 65 | -0.128 | 0.154 | 0.409 | -0.431 | 0.174 | 0.880 | 0.650 | 1.190 |
| Cholesteryl esters to total lipids ratio in small LDL | Childhood Obesity | Weighted median | 65 | -0.147 | 0.080 | 0.067 | -0.304 | 0.010 | 0.864 | 0.738 | 1.011 |
| Cholesteryl esters to total lipids ratio in small LDL | Childhood Obesity | Weighted mode | 65 | -0.146 | 0.091 | 0.115 | -0.325 | 0.033 | 0.864 | 0.723 | 1.034 |
| Omega-6 fatty acids levels | Childhood Obesity | Inverse variance weighted | 72 | -0.125 | 0.057 | 0.028 | -0.236 | -0.014 | 0.883 | 0.790 | 0.986 |
| Omega-6 fatty acids levels | Childhood Obesity | MR Egger | 72 | -0.078 | 0.100 | 0.440 | -0.274 | 0.118 | 0.925 | 0.760 | 1.126 |
| Omega-6 fatty acids levels | Childhood Obesity | Simple mode | 72 | -0.244 | 0.158 | 0.126 | -0.553 | 0.065 | 0.783 | 0.575 | 1.067 |
| Omega-6 fatty acids levels | Childhood Obesity | Weighted median | 72 | -0.148 | 0.087 | 0.088 | -0.319 | 0.022 | 0.862 | 0.727 | 1.022 |
| Omega-6 fatty acids levels | Childhood Obesity | Weighted mode | 72 | -0.173 | 0.092 | 0.065 | -0.354 | 0.008 | 0.841 | 0.702 | 1.008 |
| Phospholipids in very small VLDL | Childhood Obesity | Inverse variance weighted | 84 | -0.105 | 0.045 | 0.021 | -0.194 | -0.016 | 0.900 | 0.824 | 0.984 |
| Phospholipids in very small VLDL | Childhood Obesity | MR Egger | 84 | -0.106 | 0.070 | 0.134 | -0.242 | 0.031 | 0.900 | 0.785 | 1.032 |
| Phospholipids in very small VLDL | Childhood Obesity | Simple mode | 84 | -0.078 | 0.126 | 0.535 | -0.325 | 0.168 | 0.925 | 0.722 | 1.183 |
| Phospholipids in very small VLDL | Childhood Obesity | Weighted median | 84 | -0.111 | 0.067 | 0.096 | -0.243 | 0.020 | 0.895 | 0.785 | 1.020 |
| Phospholipids in very small VLDL | Childhood Obesity | Weighted mode | 84 | -0.140 | 0.068 | 0.042 | -0.273 | -0.007 | 0.869 | 0.761 | 0.993 |
| Ratio of 22:6 docosahexaenoic acid to total fatty acids | Childhood Obesity | Inverse variance weighted | 18 | -0.215 | 0.093 | 0.020 | -0.397 | -0.033 | 0.806 | 0.672 | 0.967 |
| Ratio of 22:6 docosahexaenoic acid to total fatty acids | Childhood Obesity | MR Egger | 18 | -0.206 | 0.138 | 0.155 | -0.476 | 0.065 | 0.814 | 0.621 | 1.067 |
| Ratio of 22:6 docosahexaenoic acid to total fatty acids | Childhood Obesity | Simple mode | 18 | -0.073 | 0.221 | 0.744 | -0.506 | 0.359 | 0.929 | 0.603 | 1.432 |
| Ratio of 22:6 docosahexaenoic acid to total fatty acids | Childhood Obesity | Weighted median | 18 | -0.185 | 0.088 | 0.035 | -0.357 | -0.013 | 0.831 | 0.700 | 0.987 |
| Ratio of 22:6 docosahexaenoic acid to total fatty acids | Childhood Obesity | Weighted mode | 18 | -0.176 | 0.081 | 0.045 | -0.336 | -0.017 | 0.838 | 0.715 | 0.983 |
| Total cholesterol levels in small HDL | Childhood Obesity | Inverse variance weighted | 34 | -0.172 | 0.084 | 0.041 | -0.337 | -0.007 | 0.842 | 0.714 | 0.993 |
| Total cholesterol levels in small HDL | Childhood Obesity | MR Egger | 34 | -0.066 | 0.132 | 0.622 | -0.325 | 0.193 | 0.936 | 0.723 | 1.213 |
| Total cholesterol levels in small HDL | Childhood Obesity | Simple mode | 34 | -0.217 | 0.196 | 0.277 | -0.602 | 0.168 | 0.805 | 0.548 | 1.183 |
| Total cholesterol levels in small HDL | Childhood Obesity | Weighted median | 34 | -0.164 | 0.117 | 0.161 | -0.393 | 0.065 | 0.849 | 0.675 | 1.068 |
| Total cholesterol levels in small HDL | Childhood Obesity | Weighted mode | 34 | -0.190 | 0.105 | 0.081 | -0.396 | 0.017 | 0.827 | 0.673 | 1.017 |
| Total cholesterol to total lipids ratio in large LDL | Childhood Obesity | Inverse variance weighted | 68 | -0.111 | 0.051 | 0.029 | -0.211 | -0.011 | 0.895 | 0.810 | 0.989 |
| Total cholesterol to total lipids ratio in large LDL | Childhood Obesity | MR Egger | 68 | -0.036 | 0.078 | 0.644 | -0.190 | 0.117 | 0.964 | 0.827 | 1.124 |
| Total cholesterol to total lipids ratio in large LDL | Childhood Obesity | Simple mode | 68 | -0.227 | 0.166 | 0.175 | -0.552 | 0.098 | 0.797 | 0.576 | 1.103 |
| Total cholesterol to total lipids ratio in large LDL | Childhood Obesity | Weighted median | 68 | -0.097 | 0.083 | 0.243 | -0.259 | 0.066 | 0.908 | 0.772 | 1.068 |
| Total cholesterol to total lipids ratio in large LDL | Childhood Obesity | Weighted mode | 68 | -0.168 | 0.105 | 0.116 | -0.374 | 0.039 | 0.846 | 0.688 | 1.040 |
| Total cholesterol to total lipids ratio in medium VLDL | Childhood Obesity | Inverse variance weighted | 47 | -0.129 | 0.060 | 0.033 | -0.247 | -0.011 | 0.879 | 0.781 | 0.989 |
| Total cholesterol to total lipids ratio in medium VLDL | Childhood Obesity | MR Egger | 47 | -0.192 | 0.107 | 0.080 | -0.402 | 0.018 | 0.825 | 0.669 | 1.019 |
| Total cholesterol to total lipids ratio in medium VLDL | Childhood Obesity | Simple mode | 47 | -0.223 | 0.143 | 0.126 | -0.504 | 0.057 | 0.800 | 0.604 | 1.059 |
| Total cholesterol to total lipids ratio in medium VLDL | Childhood Obesity | Weighted median | 47 | -0.174 | 0.083 | 0.037 | -0.338 | -0.011 | 0.840 | 0.713 | 0.989 |
| Total cholesterol to total lipids ratio in medium VLDL | Childhood Obesity | Weighted mode | 47 | -0.214 | 0.085 | 0.015 | -0.380 | -0.047 | 0.807 | 0.684 | 0.954 |
| Total cholesterol to total lipids ratio in small HDL | Childhood Obesity | Inverse variance weighted | 67 | -0.125 | 0.053 | 0.019 | -0.230 | -0.020 | 0.882 | 0.795 | 0.980 |
| Total cholesterol to total lipids ratio in small HDL | Childhood Obesity | MR Egger | 67 | -0.086 | 0.080 | 0.282 | -0.242 | 0.070 | 0.917 | 0.785 | 1.072 |
| Total cholesterol to total lipids ratio in small HDL | Childhood Obesity | Simple mode | 67 | -0.243 | 0.175 | 0.168 | -0.585 | 0.099 | 0.784 | 0.557 | 1.104 |
| Total cholesterol to total lipids ratio in small HDL | Childhood Obesity | Weighted median | 67 | -0.188 | 0.090 | 0.037 | -0.365 | -0.011 | 0.828 | 0.694 | 0.989 |
| Total cholesterol to total lipids ratio in small HDL | Childhood Obesity | Weighted mode | 67 | -0.181 | 0.095 | 0.061 | -0.367 | 0.005 | 0.834 | 0.693 | 1.005 |
| Total lipids in IDL | Childhood Obesity | Inverse variance weighted | 80 | -0.112 | 0.049 | 0.022 | -0.208 | -0.016 | 0.894 | 0.812 | 0.984 |
| Total lipids in IDL | Childhood Obesity | MR Egger | 80 | -0.127 | 0.076 | 0.102 | -0.277 | 0.023 | 0.881 | 0.758 | 1.023 |
| Total lipids in IDL | Childhood Obesity | Simple mode | 80 | -0.165 | 0.148 | 0.268 | -0.456 | 0.125 | 0.848 | 0.634 | 1.133 |
| Total lipids in IDL | Childhood Obesity | Weighted median | 80 | -0.123 | 0.074 | 0.096 | -0.269 | 0.022 | 0.884 | 0.764 | 1.022 |
| Total lipids in IDL | Childhood Obesity | Weighted mode | 80 | -0.184 | 0.084 | 0.031 | -0.349 | -0.019 | 0.832 | 0.705 | 0.981 |
| Triglycerides to total lipids ratio in medium VLDL | Childhood Obesity | Inverse variance weighted | 51 | 0.129 | 0.061 | 0.035 | 0.009 | 0.249 | 1.138 | 1.009 | 1.282 |
| Triglycerides to total lipids ratio in medium VLDL | Childhood Obesity | MR Egger | 51 | 0.167 | 0.104 | 0.115 | -0.037 | 0.371 | 1.182 | 0.964 | 1.450 |
| Triglycerides to total lipids ratio in medium VLDL | Childhood Obesity | Simple mode | 51 | 0.148 | 0.154 | 0.339 | -0.153 | 0.449 | 1.160 | 0.858 | 1.567 |
| Triglycerides to total lipids ratio in medium VLDL | Childhood Obesity | Weighted median | 51 | 0.201 | 0.086 | 0.019 | 0.033 | 0.370 | 1.223 | 1.033 | 1.448 |
| Triglycerides to total lipids ratio in medium VLDL | Childhood Obesity | Weighted mode | 51 | 0.200 | 0.082 | 0.018 | 0.040 | 0.360 | 1.222 | 1.041 | 1.434 |

| **Table S5. Results of Mendelian Randomization Analysis of Childhood Obesity on Blood Metabolites** | | | | | | | | | | | |
| --- | --- | --- | --- | --- | --- | --- | --- | --- | --- | --- | --- |
| **exposure** | **outcome** | **method** | **nsnp** | **b** | **se** | **pval** | **lo_ci** | **up_ci** | **or** | **or_lci95** | **or_uci95** |
| Childhood Obesity | Albumin levels | Inverse variance weighted | 68 | -0.005 | 0.005 | 0.331 | -0.014 | 0.005 | 0.995 | 0.986 | 1.005 |
| Childhood Obesity | Albumin levels | MR Egger | 68 | -0.003 | 0.012 | 0.782 | -0.026 | 0.019 | 0.997 | 0.974 | 1.020 |
| Childhood Obesity | Albumin levels | Simple mode | 68 | -0.019 | 0.017 | 0.274 | -0.053 | 0.015 | 0.981 | 0.949 | 1.015 |
| Childhood Obesity | Albumin levels | Weighted median | 68 | -0.008 | 0.007 | 0.277 | -0.021 | 0.006 | 0.992 | 0.979 | 1.006 |
| Childhood Obesity | Albumin levels | Weighted mode | 68 | -0.012 | 0.014 | 0.404 | -0.039 | 0.015 | 0.989 | 0.962 | 1.016 |
| Childhood Obesity | Cholesteryl esters to total lipids ratio in small HDL | Inverse variance weighted | 68 | -0.002 | 0.005 | 0.750 | -0.012 | 0.009 | 0.998 | 0.988 | 1.009 |
| Childhood Obesity | Cholesteryl esters to total lipids ratio in small HDL | MR Egger | 68 | 0.007 | 0.013 | 0.561 | -0.017 | 0.032 | 1.007 | 0.983 | 1.033 |
| Childhood Obesity | Cholesteryl esters to total lipids ratio in small HDL | Simple mode | 68 | 0.018 | 0.016 | 0.266 | -0.014 | 0.050 | 1.018 | 0.987 | 1.051 |
| Childhood Obesity | Cholesteryl esters to total lipids ratio in small HDL | Weighted median | 68 | 0.004 | 0.007 | 0.580 | -0.010 | 0.019 | 1.004 | 0.990 | 1.019 |
| Childhood Obesity | Cholesteryl esters to total lipids ratio in small HDL | Weighted mode | 68 | 0.017 | 0.013 | 0.209 | -0.009 | 0.043 | 1.017 | 0.991 | 1.044 |
| Childhood Obesity | Cholesteryl esters to total lipids ratio in small LDL | Inverse variance weighted | 68 | 0.004 | 0.005 | 0.480 | -0.006 | 0.013 | 1.004 | 0.994 | 1.014 |
| Childhood Obesity | Cholesteryl esters to total lipids ratio in small LDL | MR Egger | 68 | 0.011 | 0.012 | 0.361 | -0.013 | 0.035 | 1.011 | 0.987 | 1.036 |
| Childhood Obesity | Cholesteryl esters to total lipids ratio in small LDL | Simple mode | 68 | 0.016 | 0.017 | 0.355 | -0.017 | 0.049 | 1.016 | 0.983 | 1.050 |
| Childhood Obesity | Cholesteryl esters to total lipids ratio in small LDL | Weighted median | 68 | 0.007 | 0.007 | 0.314 | -0.007 | 0.021 | 1.007 | 0.993 | 1.021 |
| Childhood Obesity | Cholesteryl esters to total lipids ratio in small LDL | Weighted mode | 68 | 0.022 | 0.014 | 0.118 | -0.005 | 0.048 | 1.022 | 0.995 | 1.049 |
| Childhood Obesity | Omega-6 fatty acids levels | Inverse variance weighted | 68 | 0.005 | 0.005 | 0.343 | -0.005 | 0.015 | 1.005 | 0.995 | 1.015 |
| Childhood Obesity | Omega-6 fatty acids levels | MR Egger | 68 | 0.004 | 0.013 | 0.781 | -0.021 | 0.029 | 1.004 | 0.979 | 1.029 |
| Childhood Obesity | Omega-6 fatty acids levels | Simple mode | 68 | 0.005 | 0.017 | 0.772 | -0.029 | 0.039 | 1.005 | 0.971 | 1.040 |
| Childhood Obesity | Omega-6 fatty acids levels | Weighted median | 68 | 0.004 | 0.007 | 0.551 | -0.010 | 0.018 | 1.004 | 0.990 | 1.018 |
| Childhood Obesity | Omega-6 fatty acids levels | Weighted mode | 68 | 0.004 | 0.016 | 0.810 | -0.027 | 0.035 | 1.004 | 0.973 | 1.036 |
| Childhood Obesity | Phospholipids in very small VLDL | Inverse variance weighted | 68 | 0.009 | 0.005 | 0.063 | 0.000 | 0.019 | 1.009 | 1.000 | 1.019 |
| Childhood Obesity | Phospholipids in very small VLDL | MR Egger | 68 | -0.002 | 0.012 | 0.885 | -0.025 | 0.021 | 0.998 | 0.976 | 1.022 |
| Childhood Obesity | Phospholipids in very small VLDL | Simple mode | 68 | -0.004 | 0.016 | 0.829 | -0.036 | 0.028 | 0.996 | 0.965 | 1.029 |
| Childhood Obesity | Phospholipids in very small VLDL | Weighted median | 68 | 0.006 | 0.007 | 0.382 | -0.008 | 0.021 | 1.006 | 0.992 | 1.021 |
| Childhood Obesity | Phospholipids in very small VLDL | Weighted mode | 68 | 0.009 | 0.014 | 0.516 | -0.018 | 0.036 | 1.009 | 0.982 | 1.036 |
| Childhood Obesity | Ratio of 22:6 docosahexaenoic acid to total fatty acids | Inverse variance weighted | 68 | 0.001 | 0.006 | 0.920 | -0.011 | 0.013 | 1.001 | 0.989 | 1.013 |
| Childhood Obesity | Ratio of 22:6 docosahexaenoic acid to total fatty acids | MR Egger | 68 | 0.015 | 0.015 | 0.296 | -0.013 | 0.044 | 1.016 | 0.987 | 1.045 |
| Childhood Obesity | Ratio of 22:6 docosahexaenoic acid to total fatty acids | Simple mode | 68 | -0.003 | 0.018 | 0.889 | -0.038 | 0.033 | 0.997 | 0.962 | 1.034 |
| Childhood Obesity | Ratio of 22:6 docosahexaenoic acid to total fatty acids | Weighted median | 68 | 0.004 | 0.008 | 0.566 | -0.011 | 0.019 | 1.004 | 0.989 | 1.020 |
| Childhood Obesity | Ratio of 22:6 docosahexaenoic acid to total fatty acids | Weighted mode | 68 | -0.003 | 0.014 | 0.856 | -0.030 | 0.025 | 0.997 | 0.970 | 1.025 |
| Childhood Obesity | Total cholesterol levels in small HDL | Inverse variance weighted | 68 | -0.005 | 0.005 | 0.354 | -0.015 | 0.005 | 0.995 | 0.985 | 1.005 |
| Childhood Obesity | Total cholesterol levels in small HDL | MR Egger | 68 | -0.013 | 0.013 | 0.320 | -0.037 | 0.012 | 0.988 | 0.964 | 1.012 |
| Childhood Obesity | Total cholesterol levels in small HDL | Simple mode | 68 | -0.004 | 0.015 | 0.779 | -0.033 | 0.025 | 0.996 | 0.967 | 1.025 |
| Childhood Obesity | Total cholesterol levels in small HDL | Weighted median | 68 | -0.003 | 0.007 | 0.674 | -0.017 | 0.011 | 0.997 | 0.983 | 1.011 |
| Childhood Obesity | Total cholesterol levels in small HDL | Weighted mode | 68 | -0.006 | 0.012 | 0.651 | -0.030 | 0.019 | 0.994 | 0.970 | 1.019 |
| Childhood Obesity | Total cholesterol to total lipids ratio in large LDL | Inverse variance weighted | 68 | 0.002 | 0.005 | 0.663 | -0.008 | 0.012 | 1.002 | 0.992 | 1.012 |
| Childhood Obesity | Total cholesterol to total lipids ratio in large LDL | MR Egger | 68 | 0.004 | 0.012 | 0.738 | -0.020 | 0.028 | 1.004 | 0.981 | 1.028 |
| Childhood Obesity | Total cholesterol to total lipids ratio in large LDL | Simple mode | 68 | -0.016 | 0.018 | 0.362 | -0.051 | 0.019 | 0.984 | 0.950 | 1.019 |
| Childhood Obesity | Total cholesterol to total lipids ratio in large LDL | Weighted median | 68 | -0.005 | 0.008 | 0.523 | -0.020 | 0.010 | 0.995 | 0.981 | 1.010 |
| Childhood Obesity | Total cholesterol to total lipids ratio in large LDL | Weighted mode | 68 | -0.016 | 0.016 | 0.321 | -0.048 | 0.016 | 0.984 | 0.953 | 1.016 |
| Childhood Obesity | Total cholesterol to total lipids ratio in medium VLDL | Inverse variance weighted | 68 | 0.005 | 0.005 | 0.410 | -0.006 | 0.015 | 1.005 | 0.994 | 1.015 |
| Childhood Obesity | Total cholesterol to total lipids ratio in medium VLDL | MR Egger | 68 | 0.019 | 0.013 | 0.164 | -0.007 | 0.044 | 1.019 | 0.993 | 1.045 |
| Childhood Obesity | Total cholesterol to total lipids ratio in medium VLDL | Simple mode | 68 | -0.013 | 0.015 | 0.413 | -0.043 | 0.017 | 0.987 | 0.958 | 1.018 |
| Childhood Obesity | Total cholesterol to total lipids ratio in medium VLDL | Weighted median | 68 | 0.003 | 0.007 | 0.696 | -0.012 | 0.017 | 1.003 | 0.989 | 1.017 |
| Childhood Obesity | Total cholesterol to total lipids ratio in medium VLDL | Weighted mode | 68 | -0.014 | 0.014 | 0.306 | -0.041 | 0.013 | 0.986 | 0.960 | 1.013 |
| Childhood Obesity | Total cholesterol to total lipids ratio in small HDL | Inverse variance weighted | 68 | -0.004 | 0.005 | 0.401 | -0.015 | 0.006 | 0.996 | 0.986 | 1.006 |
| Childhood Obesity | Total cholesterol to total lipids ratio in small HDL | MR Egger | 68 | 0.006 | 0.012 | 0.643 | -0.019 | 0.030 | 1.006 | 0.982 | 1.031 |
| Childhood Obesity | Total cholesterol to total lipids ratio in small HDL | Simple mode | 68 | 0.015 | 0.017 | 0.372 | -0.018 | 0.049 | 1.015 | 0.982 | 1.050 |
| Childhood Obesity | Total cholesterol to total lipids ratio in small HDL | Weighted median | 68 | 0.002 | 0.007 | 0.729 | -0.011 | 0.016 | 1.002 | 0.989 | 1.016 |
| Childhood Obesity | Total cholesterol to total lipids ratio in small HDL | Weighted mode | 68 | 0.008 | 0.014 | 0.551 | -0.019 | 0.035 | 1.008 | 0.982 | 1.036 |
| Childhood Obesity | Total lipids in IDL | Inverse variance weighted | 68 | 0.007 | 0.005 | 0.139 | -0.002 | 0.017 | 1.007 | 0.998 | 1.017 |
| Childhood Obesity | Total lipids in IDL | MR Egger | 68 | -0.001 | 0.012 | 0.932 | -0.024 | 0.022 | 0.999 | 0.976 | 1.023 |
| Childhood Obesity | Total lipids in IDL | Simple mode | 68 | -0.001 | 0.017 | 0.966 | -0.034 | 0.032 | 0.999 | 0.967 | 1.033 |
| Childhood Obesity | Total lipids in IDL | Weighted median | 68 | 0.008 | 0.007 | 0.259 | -0.006 | 0.021 | 1.008 | 0.994 | 1.021 |
| Childhood Obesity | Total lipids in IDL | Weighted mode | 68 | 0.010 | 0.014 | 0.497 | -0.018 | 0.038 | 1.010 | 0.982 | 1.038 |
| Childhood Obesity | Triglycerides to total lipids ratio in medium VLDL | Inverse variance weighted | 68 | 0.003 | 0.005 | 0.637 | -0.008 | 0.013 | 1.003 | 0.992 | 1.013 |
| Childhood Obesity | Triglycerides to total lipids ratio in medium VLDL | MR Egger | 68 | -0.003 | 0.013 | 0.794 | -0.029 | 0.022 | 0.997 | 0.971 | 1.022 |
| Childhood Obesity | Triglycerides to total lipids ratio in medium VLDL | Simple mode | 68 | -0.007 | 0.019 | 0.693 | -0.044 | 0.029 | 0.993 | 0.957 | 1.029 |
| Childhood Obesity | Triglycerides to total lipids ratio in medium VLDL | Weighted median | 68 | 0.001 | 0.007 | 0.916 | -0.014 | 0.015 | 1.001 | 0.986 | 1.015 |
| Childhood Obesity | Triglycerides to total lipids ratio in medium VLDL | Weighted mode | 68 | -0.005 | 0.021 | 0.815 | -0.045 | 0.036 | 0.995 | 0.956 | 1.036 |

| **Table S6. Results of Mendelian Randomization Analysis of Blood Metabolites on Gut Microbiota** | | | | | | | | | | | |
| --- | --- | --- | --- | --- | --- | --- | --- | --- | --- | --- | --- |
| **exposure** | **outcome** | **method** | **nsnp** | **b** | **se** | **pval** | **lo_ci** | **up_ci** | **or** | **or_lci95** | **or_uci95** |
| Ratio of 22:6 docosahexaenoic acid to total fatty acids | SM23-33 | Inverse variance weighted | 18 | -0.003 | 0.018 | 0.849 | -0.039 | 0.032 | 0.997 | 0.962 | 1.033 |
| Ratio of 22:6 docosahexaenoic acid to total fatty acids | SM23-33 | MR Egger | 18 | 0.020 | 0.026 | 0.448 | -0.031 | 0.071 | 1.020 | 0.970 | 1.074 |
| Ratio of 22:6 docosahexaenoic acid to total fatty acids | SM23-33 | Simple mode | 18 | -0.056 | 0.053 | 0.304 | -0.159 | 0.047 | 0.946 | 0.853 | 1.049 |
| Ratio of 22:6 docosahexaenoic acid to total fatty acids | SM23-33 | Weighted median | 18 | 0.014 | 0.021 | 0.507 | -0.027 | 0.054 | 1.014 | 0.974 | 1.056 |
| Ratio of 22:6 docosahexaenoic acid to total fatty acids | SM23-33 | Weighted mode | 18 | 0.011 | 0.019 | 0.558 | -0.026 | 0.048 | 1.011 | 0.975 | 1.050 |
| Total cholesterol to total lipids ratio in medium VLDL | K10 sp001941205 | Inverse variance weighted | 43 | 0.013 | 0.032 | 0.689 | -0.050 | 0.075 | 1.013 | 0.952 | 1.078 |
| Total cholesterol to total lipids ratio in medium VLDL | K10 sp001941205 | MR Egger | 43 | -0.052 | 0.054 | 0.344 | -0.159 | 0.055 | 0.949 | 0.853 | 1.056 |
| Total cholesterol to total lipids ratio in medium VLDL | K10 sp001941205 | Simple mode | 43 | 0.022 | 0.073 | 0.761 | -0.121 | 0.166 | 1.023 | 0.886 | 1.180 |
| Total cholesterol to total lipids ratio in medium VLDL | K10 sp001941205 | Weighted median | 43 | 0.030 | 0.043 | 0.491 | -0.054 | 0.113 | 1.030 | 0.947 | 1.120 |
| Total cholesterol to total lipids ratio in medium VLDL | K10 sp001941205 | Weighted mode | 43 | 0.022 | 0.045 | 0.618 | -0.065 | 0.110 | 1.023 | 0.937 | 1.116 |

| **Table S7. Results of Multivariable Mendelian Randomization Analysis** | | | | | | | | | | |
| --- | --- | --- | --- | --- | --- | --- | --- | --- | --- | --- |
| **exposure** | **outcome** | **nsnp** | **b** | **se** | **pval** | **lo_ci** | **up_ci** | **or** | **or_lci95** | **or_uci95** |
| SM23-33 abundance in stool | Childhood Obesity | 0 | 0.189 | 0.142 | 0.183 | -0.089 | 0.467 | 1.208 | 0.914 | 1.596 |
| Ratio of 22:6 docosahexaenoic acid to total fatty acids | Childhood Obesity | 15 | -0.200 | 0.079 | 0.011 | -0.354 | -0.046 | 0.819 | 0.702 | 0.955 |
| K10 sp001941205 abundance in stool | Childhood Obesity | 1 | -0.074 | 0.061 | 0.222 | -0.193 | 0.045 | 0.929 | 0.825 | 1.046 |
| Total cholesterol to total lipids ratio in medium VLDL | Childhood Obesity | 38 | -0.118 | 0.059 | 0.046 | -0.234 | -0.002 | 0.889 | 0.791 | 0.998 |

| **Table S8. Sensitivity Results of Mendelian Randomization Analysis between Gut Microbiota, Blood Metabolites, and Childhood Obesity** | | | | | | | | | | | | | | | | | |
| --- | --- | --- | --- | --- | --- | --- | --- | --- | --- | --- | --- | --- | --- | --- | --- | --- | --- |
| **Exposure** | **Outcome** | **SNPs** | **MR Egger pleiotropy test** | | |  | **MR Egger Cochran’s Q heterogeneity test** | | |  | **IVW Cochran’s Q heterogeneity test** | | |  | **MR-presso text** | | |
|  |  |  | **Egger_intercept** | **SE** | **Pvalue** |  | **Q** | **Q_df** | **Q_pval** |  | **Q** | **Q_df** | **Q_pval** |  | **RSSobs** | **Pvalue** | **Outliers** |
| An181 abundance in stool | Childhood Obesity | 70 | -0.009 | 0.009 | 0.327 |  | 59.509 | 68.000 | 0.759 |  | 60.483 | 69.000 | 0.758 |  | 62.090 | 0.761 | NO |
| Bacteroides A abundance in stool | Childhood Obesity | 56 | 0.007 | 0.011 | 0.539 |  | 57.610 | 54.000 | 0.343 |  | 58.017 | 55.000 | 0.365 |  | 60.328 | 0.361 | NO |
| CAG-698 abundance in stool | Childhood Obesity | 70 | 0.001 | 0.010 | 0.944 |  | 91.186 | 68.000 | 0.032 |  | 91.193 | 69.000 | 0.038 |  | 93.554 | 0.051 | NO |
| Coprobacillus abundance in stool | Childhood Obesity | 63 | -0.010 | 0.009 | 0.293 |  | 39.715 | 61.000 | 0.984 |  | 40.841 | 62.000 | 0.983 |  | 42.280 | 0.990 | NO |
| Eubacterium I ramulus A abundance in stool | Childhood Obesity | 65 | -0.004 | 0.009 | 0.676 |  | 58.056 | 63.000 | 0.653 |  | 58.233 | 64.000 | 0.680 |  | 60.229 | 0.702 | NO |
| Faecalicatena sp001517425 abundance in stool | Childhood Obesity | 60 | 0.000 | 0.008 | 0.953 |  | 58.701 | 58.000 | 0.450 |  | 58.705 | 59.000 | 0.486 |  | 60.317 | 0.507 | NO |
| Halomonadaceae abundance in stool | Childhood Obesity | 79 | -0.009 | 0.010 | 0.401 |  | 93.750 | 77.000 | 0.094 |  | 94.616 | 78.000 | 0.097 |  | 97.098 | 0.091 | NO |
| K10 sp001941205 abundance in stool | Childhood Obesity | 68 | 0.000 | 0.009 | 0.994 |  | 51.016 | 66.000 | 0.913 |  | 51.016 | 67.000 | 0.926 |  | 52.456 | 0.925 | NO |
| Ruminococcus E sp900314705 abundance in stool | Childhood Obesity | 87 | 0.006 | 0.009 | 0.502 |  | 80.441 | 85.000 | 0.620 |  | 80.895 | 86.000 | 0.635 |  | 82.763 | 0.659 | NO |
| SM23-33 abundance in stool | Childhood Obesity | 71 | -0.001 | 0.009 | 0.941 |  | 59.614 | 69.000 | 0.783 |  | 59.619 | 70.000 | 0.807 |  | 61.365 | 0.797 | NO |
| TMED109 abundance in stool | Childhood Obesity | 64 | 0.011 | 0.009 | 0.266 |  | 51.647 | 62.000 | 0.823 |  | 52.908 | 63.000 | 0.814 |  | 54.508 | 0.831 | NO |
| UBA1206 sp000433115 abundance in stool | Childhood Obesity | 68 | 0.000 | 0.009 | 0.977 |  | 62.944 | 66.000 | 0.584 |  | 62.945 | 67.000 | 0.618 |  | 64.813 | 0.644 | NO |
| UBA8621 abundance in stool | Childhood Obesity | 66 | 0.007 | 0.009 | 0.436 |  | 58.180 | 64.000 | 0.681 |  | 58.796 | 65.000 | 0.693 |  | 60.598 | 0.701 | NO |
| Albumin levels | Childhood Obesity | 18 | -0.008 | 0.015 | 0.615 |  | 13.489 | 16.000 | 0.637 |  | 13.753 | 17.000 | 0.685 |  | 15.580 | 0.697 | NO |
| Ratio of 22:6 docosahexaenoic acid to total fatty acids | Childhood Obesity | 18 | -0.001 | 0.011 | 0.925 |  | 27.222 | 16.000 | 0.039 |  | 27.237 | 17.000 | 0.055 |  | 29.611 | 0.140 | NO |
| Omega-6 fatty acids levels | Childhood Obesity | 72 | -0.003 | 0.006 | 0.571 |  | 87.778 | 70.000 | 0.074 |  | 88.184 | 71.000 | 0.082 |  | 90.810 | 0.077 | NO |
| Total lipids in IDL | Childhood Obesity | 80 | 0.001 | 0.005 | 0.802 |  | 94.701 | 78.000 | 0.096 |  | 94.778 | 79.000 | 0.109 |  | 98.531 | 0.088 | NO |
| Total cholesterol to total lipids ratio in large LDL | Childhood Obesity | 68 | -0.006 | 0.005 | 0.215 |  | 71.490 | 66.000 | 0.301 |  | 73.186 | 67.000 | 0.282 |  | 77.392 | 0.243 | NO |
| Total cholesterol to total lipids ratio in medium VLDL | Childhood Obesity | 47 | 0.005 | 0.007 | 0.481 |  | 50.519 | 45.000 | 0.265 |  | 51.087 | 46.000 | 0.281 |  | 53.013 | 0.319 | NO |
| Triglycerides to total lipids ratio in medium VLDL | Childhood Obesity | 51 | -0.003 | 0.007 | 0.651 |  | 59.295 | 49.000 | 0.149 |  | 59.546 | 50.000 | 0.167 |  | 61.572 | 0.174 | NO |
| Total cholesterol levels in small HDL | Childhood Obesity | 34 | -0.008 | 0.007 | 0.307 |  | 36.571 | 32.000 | 0.265 |  | 37.804 | 33.000 | 0.259 |  | 39.192 | 0.305 | NO |
| Total cholesterol to total lipids ratio in small HDL | Childhood Obesity | 67 | -0.003 | 0.005 | 0.512 |  | 65.423 | 65.000 | 0.462 |  | 65.861 | 66.000 | 0.482 |  | 69.445 | 0.466 | NO |
| Cholesteryl esters to total lipids ratio in small HDL | Childhood Obesity | 64 | -0.003 | 0.005 | 0.531 |  | 63.396 | 62.000 | 0.427 |  | 63.802 | 63.000 | 0.448 |  | 66.972 | 0.416 | NO |
| Cholesteryl esters to total lipids ratio in small LDL | Childhood Obesity | 65 | 0.000 | 0.005 | 0.935 |  | 57.705 | 63.000 | 0.665 |  | 57.712 | 64.000 | 0.697 |  | 61.133 | 0.641 | NO |
| Phospholipids in very small VLDL | Childhood Obesity | 84 | 0.000 | 0.004 | 0.987 |  | 92.500 | 82.000 | 0.201 |  | 92.500 | 83.000 | 0.223 |  | 95.343 | 0.212 | NO |
| SM23-33 abundance in stool | Ratio of 22:6 docosahexaenoic acid to total fatty acids | 71 | -0.002 | 0.002 | 0.281 |  | 71.782 | 69.000 | 0.386 |  | 73.009 | 70.000 | 0.379 |  | 75.229 | 0.378 | NO |
| K10 sp001941205 abundance in stool | Total cholesterol to total lipids ratio in medium VLDL | 70 | 0.001 | 0.002 | 0.522 |  | 67.371 | 68.000 | 0.499 |  | 67.785 | 69.000 | 0.519 |  | 69.600 | 0.547 | NO |
